# Development of the Technology Readiness Inventory for Professionals (TRIP): field testing in Dutch addiction care professionals

**DOI:** 10.1101/2025.11.04.25339119

**Authors:** Ruben Veltman, Joanne E.L. VanDerNagel, Marloes G. Postel, Peter M. ten Klooster, Marjolein E.M. Den Ouden, Saskia M. Kelders

**Author notes:** Corresponding author E-Mail (RV).

## Abstract

For the successful adoption of technology in healthcare, professionals need to possess or develop a range of technology competencies, attitudes and resources. However, there is a lack of feasible, valid and reliable questionnaires to assess these attributes.

An existing item set of 90 items across nineteen subscales, originally developed to measure adoption and technology competencies in health education, was adapted for healthcare professionals and administered in a cross-sectional online survey among addiction care professionals (*n* = 334) in the Netherlands.

Item analysis and exploratory factor analysis (EFA) were used to reduce the number of items and to determine the underlying factor structure of the inventory. EFA suggested that the original nineteen hypothesised subscales tapped into three reliable underlying factors: (1) technology competencies (ω = 0.94), (2) attitude towards the use of technology (ω = 0.93) and (3) facilitating conditions (ω = 0.89). The Pearson intercorrelations were *r* = 0.53 (*p* < 0.001) between factors 1 and 3, *r* = 0.47 (*p* < 0.001) between factors 1 and 2 and *r* = 0.36 (*p* < 0.001) between factors 2 and 3, indicating that the questionnaire measures related, but sufficiently distinct underlying constructs supporting their discriminant validity.

The 90 items in the original item set were reduced to 49 items that together provide a psychometrically sound and feasible inventory for measuring technology competencies, attitude and facilitating conditions in an addiction care context. Further research is needed to thoroughly validate the questionnaire, also in different healthcare settings and to confirm its dimensionality, reliability, convergent and discriminant validity, and criterion validity.

**Author Summary:** Health related technologies (eHealth), such as online treatment platforms, can contribute to empowering people and improving accessibility and quality of care. To be able to use eHealth, healthcare professionals should adopt (health) technology and have technology competencies. In our study we focus on addiction care workers. Gaining insight into the adoption readiness and technology competencies of professionals working in addiction care is important to better understand what is needed to implement eHealth in this setting. We adapted an existing item set, originally developed to measure technology competencies and adoption in educational settings, to the context of addiction care. Furthermore, we aimed to validate and shorten this item set to a new inventory, suitable to apply in addiction care settings. Psychometric analyses of responses of 334 addiction care professional resulted in a 49-item Technology Readiness Inventory for Professionals (TRIP) measuring (1) technology competencies, (2) attitude toward technology and (3) facilitating conditions needed to be able to use technology. More research is needed to further validate TRIP, to apply it in different contexts and to determine whether it can also measure changes over time within an organisation.

## Introduction

Using eHealth in healthcare provides a strategy to address the challenges of staff shortages and increasing healthcare costs [1,2]. eHealth can be defined as “the use of technology to support health, well-being and healthcare” [3 p6]. This broad definition entails different types of technology including telehealth (videoconferencing between patient and treatment provider), online therapy (e.g. internet delivered cognitive behavioural therapy) and virtual reality (e.g. for treating addiction, depression, anxiety, etc.). eHealth contributes to empowering people and can improve accessibility and quality of care [3]. Despite these benefits, the use of eHealth in healthcare is not yet standard clinical practice.

The lack of eHealth uptake and integration can be attributed to several factors, including the inherent complexity, unpredictability, and nonlinearity of healthcare systems [4,5], limited availability of resources [6], and insufficient technological adoption and digital competencies among healthcare professionals [7]. The latter (adoption and competencies) are key elements in successfully implementing health care technology [8] and are the main focus of this study.

Both constructs have not yet a complete and fully accepted definition. However, in this study adoption entails the decision of end users to begin using new technologies [9] and competencies entail knowledge of and attitude towards the use of eHealth [10], and a “mixture of digital literacy, critical reflexion, problem solving skills and confidence in technology” [11 p22].

Gaining insight into the adoption readiness and technology competencies of healthcare professionals is important to better understand what is needed to implement eHealth in this setting. To date, however, there are no validated questionnaires available that are suitable for the healthcare context and that capture the full scope of both adoption and technological competencies.

In the context of healthcare and wellbeing education, however, a set of 90 items measuring teachers’ adoption readiness and technology competencies was recently developed in the Netherlands [11], following the steps of the AMEE Guide of questionnaire development [12]: (1) literature review, (2) focus group interviews, (3) synthesising data of literature review and focus groups, (4) item development, (5) expert validation and refinement of items and (6) cognitive interviews with potential respondents to test the comprehensibility of the items and (7) pilot testing. In this item set, adoption is reflected by the following constructs: attitude towards technology, attitude towards educating colleagues (in applying technology), self-efficacy, external influence, behavioural intention, facilitating conditions and knowledge of and experience with technology (see S1 Table for all items and their sources). These constructs were synthesised from different models (step 3), such as the (extended) Unified Theory of Acceptance and Use of Technology (UTAUT) [13,14] and the Technology Readiness and Acceptance Model (TRAM) [15,16], and expert consultation (step 5).

In the item set, competencies are divided in basic and advanced competencies, in accordance with the Dutch technology competency model [17,18] (see S2 Table for an overview). According to this model, every professional in healthcare should possess a set of basic technology competencies (e.g. knowing how to operate a digital patient portal). Advanced technology competencies (e.g. informing your colleagues and organisation about technology) are expected of professionals who take a leading role in technology within their organisation. Constructs of basic technology competencies are changing (care through technology), finding technology, being confident in using technology, skilfully using technology and instructing your client about technology. Constructs of advanced technology competencies are leading, deepening, connecting, informing, improving, and replacing. Ethical reflection is an overarching skill in the model. These constructs were also synthesised from different models (step 3), such as the Digital Literacy Scale (DLS) [19] and students’ digital competence scale (SDiCoS) [20], and expert consultation (step 5).

This study aims to adapt the item set from the context of health and wellbeing education to the context of healthcare and to develop a feasible (i.e. brief and comprehensible), reliable and valid self-report measure of technology adoption and competencies. For this study, we selected the addiction care context to develop and evaluate the measure, given the significant role that digitisation plays in this field.

## Method

### Ethics statement

This study was conducted at the institute of Tactus Addiction Care in the eastern part of the Netherlands and was approved by the ethical committee of the University of Twente in May 2023 (reference number 230334) and the Scientific Committee of Tactus Addiction Care in June 2023.

Participants were asked to complete the online survey in Qualtrics after reading the study information and providing informed consent.

### Design

Following the design of a cross-sectional online survey study, we field tested the adoption readiness and technology competencies item set among a convenience sample of care providers in a Dutch addiction care institute, with the aim of developing a feasible and psychometrically sound inventory for application in various care settings. Field testing aligns with step 7 of the AMEE Guide for developing questionnaires, which emphasizes the importance of preliminary testing to evaluate item quality and refine the instrument [12].

### Participants

Potential participants were treatment providers (*N* = 998) working in the primary care process (e.g. social workers, psychologists, nurses, psychiatrists, etc.) of Tactus addiction care. Tactus addiction care has over 1,600 employees, working in 46 different locations of care, cure and assisted living, treating thousands of patients [21].

### Measurements

The item pool (see S1 Table) consisted of 90 items with a 5-point Likert-type response scale format (ranging from strongly disagree to strongly agree), divided over 19 subscales. These subscales were rationally constructed based on theoretical models and existing questionnaires, such as the unified theory of acceptance and use of technology (UTAUT) [13], extended UTAUT [14], technology readiness and acceptance model (TRAM) [15,16], Dinamo 6.0 [22] and expert insights. Technology competencies were derived from the Dutch technology competency model [17], refined by Den Ouden et al. [18].

Adoption subscales were (1) attitude towards technology (AT), (2) attitude towards educating colleagues (in applying technology) (AE), (3) self-efficacy (SE), (4) external influence (EI), (5) behavioural intention (BI), (6) facilitating conditions (FC), and (7) knowledge of and experience with technology (KET). Subscales of five basic competencies in using technology were (8) changing (CHA), (9) finding (FIN), (10) being confident (COF), (11) skilfully using technology (SU), and (12) instructing (INS). Lastly, subscales of six advanced technology competencies were (13) leading (LEA), (14) deepening (DEE), (15) connecting (CON), (16) informing (INF), (17) improving (IP), (18) replacing (REP), and (19) an overarching skill of ethical reflection (ER).

With expert consultation between the co-authors (MP and JvdN) and the first author (RV) the version for the (health and wellbeing) educational context was transposed to a new inventory of items suitable for addiction care professionals without changing the meaning and number of items. Before administering the inventory, three experts (two researchers and an eHealth specialist) provided feedback on the adaptation of the items from the educational context to addiction care, as well as on the wording and flow of the items and instructions. Based on this, the instruction text was shortened and the wording of some items was adjusted to increase comprehensibility.

### Procedure

Participants were recruited via their institution email addresses after being informed by their managers. They completed the online survey in Qualtrics, which took approximately 20 to 30 minutes.

To increase the response rate, managers were asked to motivate their team members, responders could win a cake for their entire team and initial non-responders received a maximum of six reminders.

### Data Analysis

Data were analysed using the Statistical Package for the Social Sciences (SPSS) version 29 [23]. Discontinued questionnaires with non-random missing values (*n* = 53) were excluded from analysis.

In several iterations, item reduction analyses were performed to decrease the number of items. Results of each iterative step were discussed within the team of authors.

First, we performed an additional examination of the content of the items and removed those that were negatively, ambiguously, or vaguely formulated (e.g., “Technology lowers the quality of relationships by reducing personal interaction”). Negatively formulated items can increase measurement error [24], and can cause misinterpretations and method effects (creating artificial factors for positively and negatively worded items) [25,26].

Second, Artino et al. [12] recommend reviewing the distribution of the data, such as variance. In our study items with relatively high variances were considered beneficial, as they are better capable of distinguishing individual differences in competence and adoption levels between people. Consequently, items with a relatively low variance (< 0.6), were discarded. This threshold score represents 15% of the maximum possible variance in our dataset.

Third, item homogeneity was gauged by calculating the inter-item-correlation with Spearman’s rho within the entire dataset. Item-pairs that correlated very highly (*r*_*s*_ > .7), pointing to item redundancy, were then discussed within the team (RV, JvdN, MP, SK) based on their content. Of each redundant pair, the item with the most clear wording was then retained.

Four, low correlating item-pairs (*r*_*s*_ < .4) within assumed subscales were discussed between the authors, because low correlating item-pairs don’t both fit within the same subscale. For each item pair, the item with most concise wording was retained.

Five, Cronbach’s alpha and item-rest correlations were computed within the assumed subscales. Items exhibiting a low item-rest correlation (*r* < 0.4) that also had an ambiguous content, and whose removal yielded a positive contribution to the alpha value, were discarded. After discarding items in a subscale, several iterations of this analysis were performed until there were no more items with low item-rest correlations.

The remaining items were subjected to a series of exploratory factor analyses with principal axis factoring. An oblique rotation (direct oblimin) was conducted, assuming that there would be some degree of correlation between latent factors. The Kayser-Meyer-Olkin (KMO) measure was performed to examine sampling adequacy based on a minimum criterion of 0.5 [27]. In addition, Bartlett’s test of sphericity was performed to determine whether the correlation matrix was significantly different from an identity matrix. If the associated chi-square test proved to be significant, data were considered sufficiently correlated to perform a factor analysis. Items with factor loadings under 0.3 were left out in subsequent EFA iterations. Cross loadings between factors were examined to assess the distinctiveness of the constructs and to identify potential overlap between items across factors. The number of factors to retain was based on Cattell’s screeplot [28]. After discussion between authors, more items were discarded in the final iteration of the EFA because their content did not sufficiently represent the label of the factor on which they loaded. Subsequently, McDonald’s omega was calculated to test the composite reliability of each final latent variable (factor). Values between 0.70 and 0.95 were considered indicative of good internal consistency [29].

## Results

### Study Sample

The survey was sent to all 998 treatment providers in the addiction care institute. The number of participants that completed the questionnaire was 334 (response rate 33.47%), including 245 (73.4%) women and 85 men (25.4%) and four persons who did not want to disclose their gender, with a mean age of 40.4 (*SD* = 12.0) and a range of 21 to 68 years old (see Table 1 for demographics).

**Table 1.** Socio-demographic characteristics of the study population (*N* = 334)

| **Variable** |  |
| --- | --- |
| Age (years) (n = 331) | Mean (SD) |
|  | 40.4 (12.0) |
| Gender | n (%) |
| Woman | 245 (73.4) |
| Man | 85 (25.4) |
| Not disclosed | 4 (1.2) |
| Profession | n (%) |
| Social Worker | 90 (26.9) |
| Nurse | 66 (19.8) |
| Psychologist | 46 (13.8) |
| Psychiatrist | 9 (2.7) |
| Psychomotor therapist | 5 (1.5) |
| (Addiction) Physician | 3 (0.9) |
| Other profession | 115 (34.4) |
| Professional Experience with technology | n (%) |
| < 1 year | 59 (17.7) |
| 1 to 4 years | 193 (57.8) |
| 5 to 9 years | 52 (15.6) |
| ≥ 10 years | 30 (9) |

### Item reduction analyses and internal construct validation

All steps taken to reduce the number of items and to retain a feasible, structurally valid and reliable new questionnaire are depicted in Fig 1 (see S1 Table for a listing of all 90 items and specific reasons for the 41 items that were removed). In step one, eight negatively formulated items were removed. Next, four items were discarded based on their relatively small variance (< 0.6) and elusive meaning. Four additional items were removed due to homogeneity (*r*_*s*_ > 0.7). One of those items correlated lower than 0.7 with another item (*r*_*s*_ = 0.65), but was removed nevertheless due to the very similar meaning of the two items. Two items were discarded based on a low inter-item correlation (*r*_*s*_ < 0.4) with other items within the assumed same subscale in combination with elusive item content. Two items with low item-rest correlation (*r*_*s*_ < 0.4) and which negatively affected Cronbach’s alpha within a subscale, were removed. After removal, Cronbach’s alphas and item-rest correlations in the remaining seventy items showed no indications to discard more items at this point.

**Fig 1.**
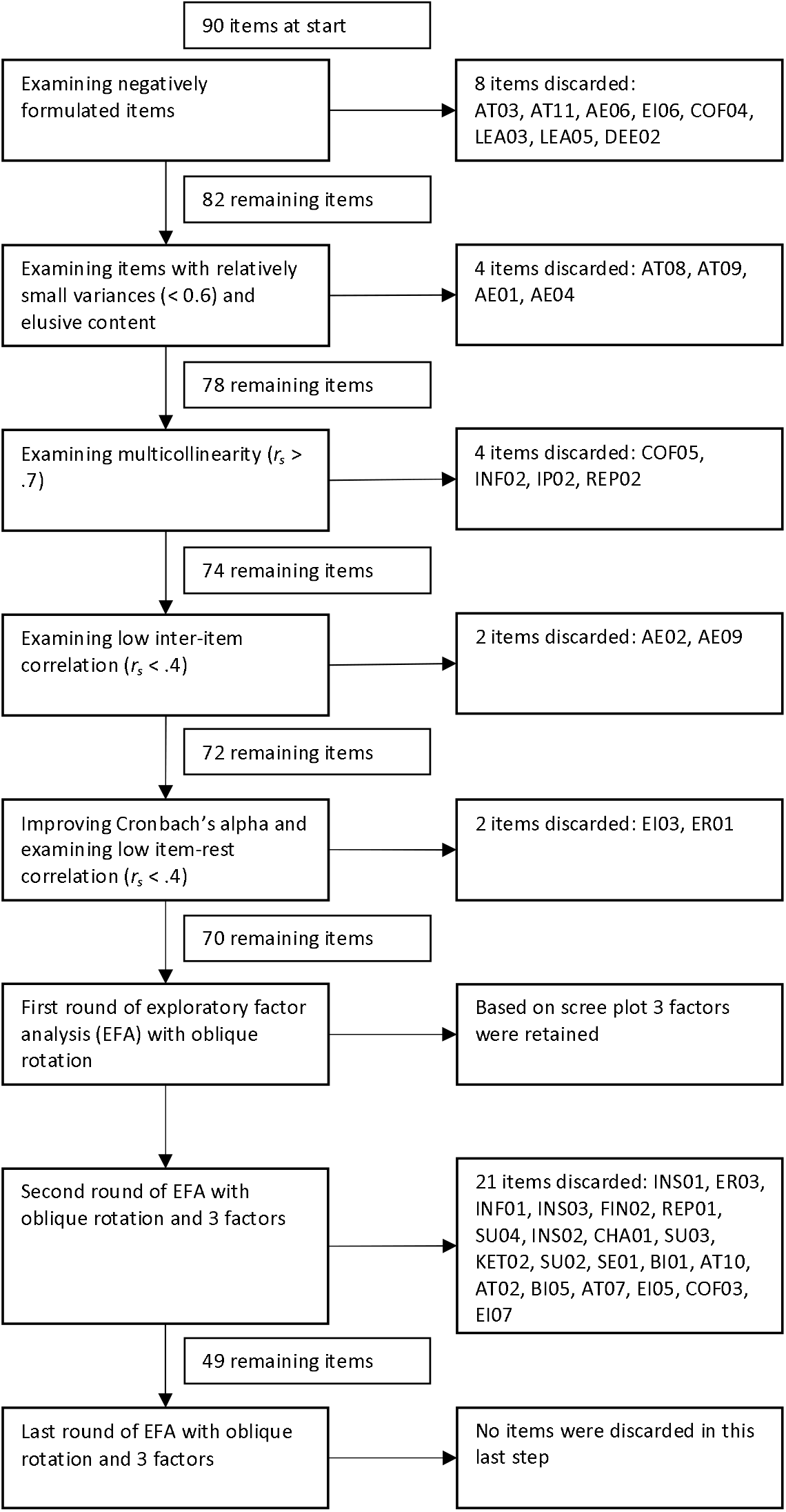
Steps in item reduction analyses.

To examine the structural validity of the questionnaire, EFA was executed on the remaining seventy items. The Kayser-Meyer-Olkin (KMO) measure of sampling adequacy was 0.94, suggesting that factor analysis should yield distinct and reliable factors. KMO-values of all individual items were greater than 0.88 and most of them above 0.90. Bartlett’s test of sphericity was significant (χ^2^(1176) = 9948.57, p < 0.001), suggesting that items correlated sufficiently to perform a factor analysis. An initial EFA was executed to decide how many factors to retain. Based on Catell’s screeplot three factors were retained that explained 47.74% of the variance.

A second iteration of EFAs was executed with a fixed number of three factors. The items that clustered on the factors suggested that factor 1 mainly included items measuring technology competencies, factor 2 attitude towards use of technology and factor 3 facilitating conditions necessary to be able to use technology. However, the content of 21 items did not sufficiently match the label of the latent variable (factor) on which they loaded. Consequently, they were removed. Three of these 21 items also had substantial cross loadings on two factors (> 0.3) (i.e. SU02, SE01 and EI05).

With the remaining 49 items a third and last iteration of EFA was executed. Table 2 shows EFA results of this last iteration after an oblique rotation. This factor model demonstrated a clear solution with all items loading substantially (≥ 0.40) on their respective factor. Eigenvalues were 15.27, 4.71 and 3.82, respectively and the three factors together explained 48.58 % of the variance in item responses.

**Table 2.** Factor loadings on three factor solution containing 49 items (*N* = 334)

|  | Rotated Factor Loadings<sup>a</sup> |  |  |
| --- | --- | --- | --- |
|  | 1 Technology competencies | 2 Attitude towards use of technology | 3 Facilitating conditions |
| I have expertise in implementing (new) technology (LEA04) | 0.80 |  |  |
| Colleagues and clients come to me for advice and questions about (new) technology (LEA01) | 0.78 |  |  |
| I can explain the impact of technology to clients/colleagues on their work (INS05) | 0.71 |  |  |
| My supervisor asks me for advice on (new) technological developments (LEA02) | 0.71 |  |  |
| I identify the needs and requirements of colleagues and clients regarding technology knowledge and skills (CON01) | 0.67 |  |  |
| I advise my organisation solicited and unsolicited on the use of (new) technology by clients/colleagues (CON04) | 0.67 |  |  |
| I can support clients/colleagues in selecting technology for an individual request for help (SU06) | 0.66 |  |  |
| I can tailor the information I give to what clients/colleagues need to apply technology (INS04) | 0.66 |  |  |
| I inspire clients/colleagues to use technology (INF03) | 0.65 |  |  |
|  | 1 Technology competencies | 2 Attitude towards use of technology | 3 Facilitating conditions |
| I have ideas on how to improve the use of technology in treatment and counselling (IP01) | 0.64 |  |  |
| I actively seek substantiation to deploy a (new) technology (DEE01) | 0.64 |  |  |
| I discuss ethical dilemmas around the use of technology with my clients/colleagues (ER02) | 0.62 |  |  |
| I can support clients/colleagues in applying data protection rules when using technology (SU05) | 0.61 |  |  |
| I can (technically) support clients/colleagues in using technology (SU01) | 0.60 |  |  |
| Through my connections with the other professionals/suppliers in the practice, I remain well informed about technology used there (CON03) | 0.60 |  |  |
| I encourage clients/colleagues to be alert to the safe use of technology (INF04) | 0.57 |  |  |
| I can assess whether technological innovations are appropriate for my professional practice (DEE03) | 0.57 |  |  |
| I know how to find reliable information about technology in addiction care (FIN01) | 0.54 |  |  |
| I interact with other professionals in the field who use technology in their work (CON02) | 0.53 |  |  |
| I possess knowledge needed to apply technology in my work (KET01) | 0.51 |  |  |
|  | 1 Technology competencies | 2 Attitude towards use of technology | 3 Facilitating conditions |
| I believe in the added value of technology within addiction care (COF01) |  | -0.80 |  |
| I find it important to learn about technology (AE07) |  | -0.76 |  |
| I am curious about developments in technology for addiction care (BI04) |  | -0.73 |  |
| Technology is necessary to future-proof addiction care (AT06) |  | -0.72 |  |
| I am confident that I can learn to apply technology in my work (SE02) |  | -0.70 |  |
| Technology makes it easier for me to do my job (AT13) |  | -0.67 |  |
| I am willing to (continue to) develop in the field of technology (BI02) |  | -0.67 |  |
| Technology should be a natural part of the training offer for addiction care professionals (AE08) |  | -0.66 |  |
| Technology adds value for my clients (AT12) |  | -0.65 |  |
| Technology leads to a higher quality of my work (AT04) |  | -0.63 |  |
| I am willing to review my role as an addiction treatment professional in the context of technology developments (BI03) |  | -0.62 |  |
|  | 1 Technology competencies | 2 Attitude towards use of technology | 3 Facilitating conditions |
| Technology gives me more job satisfaction (AT01) |  | -0.58 |  |
| Technology contributes to more self-reliance for my clients (AT14) |  | -0.57 |  |
| I am willing to discuss resistance to technology among colleagues (BI06) |  | -0.55 |  |
| Technology leads to time savings for me (AT05) |  | -0.54 |  |
| Developments in the field require me to have knowledge and skills in technology (EI01) |  | -0.53 |  |
| I am willing to support colleagues in working with technology (AE03) |  | -0.50 |  |
| I am confident in my own abilities regarding the use of technology in addiction care (COF02) |  | -0.48 |  |
| I find it important to engage with colleagues about the possibilities and limitations of technology (AE05) |  | -0.44 |  |
| My organisation provides funding to train employees in the use of technology (FC04) |  |  | 0.81 |
| I am facilitated in attending courses or training on technological developments in addiction care (FC02) |  |  | 0.79 |
| My organisation provides sufficient time to learn to use technology (FC03) |  |  | 0.78 |
| I have easy access to technologies used within addiction care (FC05) |  |  | 0.76 |
|  | 1 Technology competencies | 2 Attitude towards use of technology | 3 Facilitating conditions |
| My organisation provides innovation space to experiment with technology in addiction care (FC06) |  |  | 0.70 |
| My organisation has a clear vision regarding technology in addiction care (FC01) |  |  | 0.62 |
| My supervisor encourages a focus on technology in helping clients (EI02) |  |  | 0.57 |
| My supervisor expects me to have sufficient knowledge of technology to prepare clients and colleagues to use it (EI04) |  |  | 0.48 |
| Technology is a topic of conversation within my organisation (EI09) |  |  | 0.46 |
| Colleagues find it important that technology is part of their work (EI08) |  |  | 0.40 |
| Eigenvalues | 15.27 | 4.71 | 3.82 |
| % of variance | 31.17 | 9.60 | 7.80 |

<sup>a</sup>Factor loadings < 0.3 were not included in the table.

Table 3 presents the composite reliability coefficients, mean scores and Pearson intercorrelations of the observed subscales. The three factors demonstrated good internal consistency, suggesting that the item responses in each factor can be averaged to a reliable subscale score. Intercorrelations between the subscale scores were moderate to strong, indicating that the questionnaire measures related, but sufficiently distinct underlying constructs supporting their divergent validity.

**Table 3.**
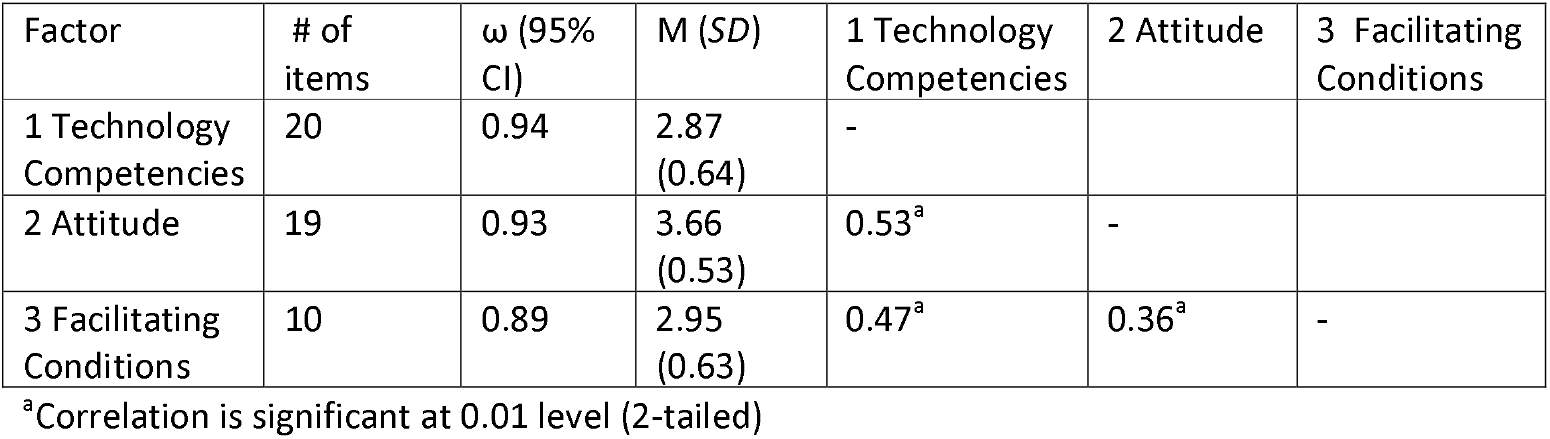
Internal consistency, mean scores and Pearson intercorrelations of the observed factors (*N* = 334)

| Factor | # of items | $\omega$ (95% CI) | M (SD) | 1 Technology Competencies | 2 Attitude | 3 Facilitating Conditions |
| --- | --- | --- | --- | --- | --- | --- |
| 1 Technology Competencies | 20 | 0.94 | 2.87 (0.64) | - |  |  |
| 2 Attitude | 19 | 0.93 | 3.66 (0.53) | 0.53<sup>a</sup> | - |  |
| 3 Facilitating Conditions | 10 | 0.89 | 2.95 (0.63) | 0.47<sup>a</sup> | 0.36<sup>a</sup> | - |

<sup>a</sup>Correlation is significant at 0.01 level (2-tailed)

## Discussion

In this study, we aimed to develop a feasible (i.e. brief and comprehensible), internally consistent, and valid self-report measure of technology adoption and competencies in addiction care, based on a comprehensive item set originally developed for health education contexts. The analyses contributed to a first step towards a valid, reliable and compact questionnaire of 49 items measuring technology competencies, attitude and facilitating conditions needed to apply technology in an addiction care context. The original item set was derived from a comprehensive collection of models and frameworks that measure comparable concepts, resulting in many subscales. Results show that after exploratory factor analysis (EFA), the original nineteen hypothesised subscales can be reduced to three underlying factors: (1) technology competencies, (2) attitude towards the use of technology and (3) facilitating conditions. This suggests that the original rationally derived conceptualisation may have assumed an overly differentiated depiction of the underlying structure, which is better represented empirically by three broader factors. Even though items derive from different models, they appear to measure the same constructs. By reducing the number of items from 90 to 49 and by exploring which latent constructs can be clearly distinguished, a new questionnaire, coined the Technology Readiness Inventory for Professionals (TRIP), has been created that can be used in an addiction care context. Per construct mean scores and standard deviations can be calculated to gain insight in technology competencies, attitude and facilitating conditions. Scores range from 1 to 5 and the higher the score, the more positively the given construct is rated. Further research is required to establish normative values and to determine how score differences relate to meaningful outcomes.

The first factor of technology competencies underlying the final items implies that the presumed distinction between basic and advanced technology competencies was not supported by the data; they actually tap into one and the same dimension. A likely explanation is that participants that score highly on advanced competencies also score highly on basic competencies and similarly for low scores. This indicates that basic and advanced technology competencies may be perceived as a continuum rather than as distinct categories. Interestingly, the items of the technology competency being confident (in using technology) (COF) was not retained within the construct of technology competencies. Based on individual factor loadings, this construct appeared more closely related to attitude toward technology instead of competencies. This is consistent with the conceptual overlap between the self-efficacy item, which includes the term confident and loaded on the attitude factor. Furthermore, the competencies changing (CHA) and replacing (REP) were not included in any of the three identified factors. Both competencies were derived through expert consultation rather than being grounded in theoretical frameworks. Previous studies have shown that theory-based items often demonstrate stronger psychometric properties [30]. As such, expert-derived items may be more susceptible to revision or exclusion. In addition, they don’t appear to have a unique contribution to technology competencies, but seem to be broader concepts. They appear to be rather multidimensional, including knowledge, influence, communication, and reflective and societal insights.

The second factor was labelled as attitude. This label seems more suitable than the hypothesised construct of adoption in the original item set, because there is a substantive connection in the content of the items that represent attitude: the items include beliefs, feelings of competence, willingness to take action, and appreciation for technological developments. This partly corresponds with the definition of attitude in the Theory of Planned Behavior (TPB), that entails the extent to which an individual has a positive or negative assessment or opinion of the target behaviour (using technology) [31]. In the TPB attitude is seen as one of the main determinants of behavioural intention, besides subjective norm and perceived behavioural control. The original construct (adoption), and many of the items were based on or derived from the field of technology adoption, including models as the Technology Acceptance Model (TAM) [32] and Unified Theory of the Use and Acceptance of Technology (UTAUT) [13]. In TAM, attitude is seen as a mediator of perceived usefulness and perceived ease of use that leads to behavioural intention. However, in later technology acceptance models, attitude is no longer a construct that influences behavioural intention and the actual use of technology. This is illustrated in the UTAUT [13] that shows that attitude has a spurious relation with behavioural intention when performance expectancy and effort expectancy were added as important predictors of behavioural intention. Psychometrically, the second factor measures a single construct. Theoretically, and considering the content of the items in the second factor, there are three other underlying constructs that could be distinguished in accordance with those defined in UTAUT: performance expectancy (items COF01, AT06, AT13, AT12, AT04, AT14, AT05, AT01, EI01), behavioural intention (items BI02, BI03, BI04, BI06, AE03, AE05, AE08, AE07) and effort expectancy (items SE02, COF02). Empirically, however, these three constructs appear to be strongly interrelated and may therefore represent manifestations of a single underlying construct. See S1 Table for the content of these items. Future research in new samples is needed to confirm or reject the hypothesis that there might be three underlying constructs, e.g. by performing confirmative factor analysis in a new sample that completes this newly developed TRIP questionnaire.

The third factor measures facilitating conditions needed to be able to use technology. This factor consists of several items of the constructs facilitating conditions (FC) and external influence (EI) of the original item set. Items of facilitating conditions derived from Extended UTAUT [14] and predict behavioural intentions, next to effort and performance expectancy [13,33]. Items of external influence derive from UTAUT [13] and Dinamo 6.0 [22] (a questionnaire measuring willingness to change of professionals within an organisation) and expert consultation, and appear to correspond with the construct of social influence from UTAUT. The existence or absence of these underlying constructs in this factor (FC and EI) should be scrutinised in future research. If the existence of these underlying constructs is confirmed, factors 2 and 3 may be representations of the constructs performance expectancy, effort expectancy, social influence and facilitating conditions, predictors influencing behavioural intention in UTAUT. If that is the case, both factors might be different representations of acceptance of technology as defined in the UTAUT model. However, future research is needed to confirm the dimensions underlying the TRIP and their reliability, and convergent, discriminant and criterion validity [34].

In this study we field tested the TRIP questionnaire, aligning with step 7 of the AMEE Guide for developing questionnaires, which emphasizes the importance of preliminary testing to evaluate item quality and refine the instrument [12]. However, more research is needed to get insight into the feasibility of the questionnaire and how it can be applied in the addiction care practice. Scrutiny is needed to determine if TRIP can indeed measure diverse patterns and nuances in technology competencies, attitudes and facilitating conditions and whether it can be effectively applied across various (health)care contexts. Furthermore, it is worthwhile to investigate the potential of TRIP to help mitigate barriers to technology adoption, thereby enhancing uptake. Finally, it is important to examine if the TRIP questionnaire is suitable for measuring organisational change over time, enabling longitudinal monitoring.

### Limitations

The relatively low response rate (33.47 %) is one of the limitations of the current study. A possible reason for this might be the length of the original item set, that made completing the survey burdensome for participants. Other reasons might be the high workload of the target group and their priority to client work. Although data of non-responders were lacking, and the sample might not have been representative for the entire population, the sample size appeared adequate for examining the psychometric properties of the item set.

Second, our sample consisted mostly of social workers, nurses and psychologists. This is unsurprising, as they present the largest professional groups in our population. Other professional groups (e.g. psychiatrists, psychomotor therapists, physicians) were less represented in our sample. Over or under representativeness may cause a sampling bias [35]. However, for examining the psychometric properties of the item set, representativeness was considered less relevant than sample size, because we didn’t expect psychometric properties to be different in other professions.

Our sample of 334 participants did not meet the common rule of thumb requirement of at least ten participants per item for robust factor analysis [36]. However, according to Field [37], the total sample size, factor loadings and communalities are more important. Other studies have considered a sample size of 300 or more as good (37) to give a stable factor solution. It remains a challenge to recruit addiction care professionals for non-care-related tasks in current times of staff shortages and high workloads.

Lastly, as our questions did not specify a specific type of technology, participants may have had different technologies in mind which may have influenced their answers. A reason for not specifying a specific type of technology was that TRIP aims to capture insights into technology-related competencies, attitudes, and facilitating conditions across a range of technologies. Therefore, this wider focus was intentionally chosen in order to measure these constructs in a broader perspective, given the fact that different types of technology are already available within the addiction care context.

## Conclusion

In this study, a first step has been taken to develop TRIP, a psychometrically sound and feasible questionnaire for measuring technology competencies, attitude toward technology and facilitating conditions to be able to use technology for professionals working in an addiction care setting. A recommendation for future research is to perform a confirmatory factor analysis in a new sample to examine if there might be other underlying (sub)constructs, to reduce more items, and testing for dimensionality, reliability, and convergent, criterion and discriminant validity. Furthermore, future research should also focus on the feasibility for application in addiction care and other healthcare settings.

## Data Availability

Data cannot be shared publicly because of sensitive human research participant data. Data are available upon request from the Scientific Committee of the Institution of Tactus Addiction Care, for researchers who meet the criteria for access to confidential data. Head of the Scientific Committee dr. Jeannette van Manen, can be contacted via.

## Supporting information

**S1 Table.**
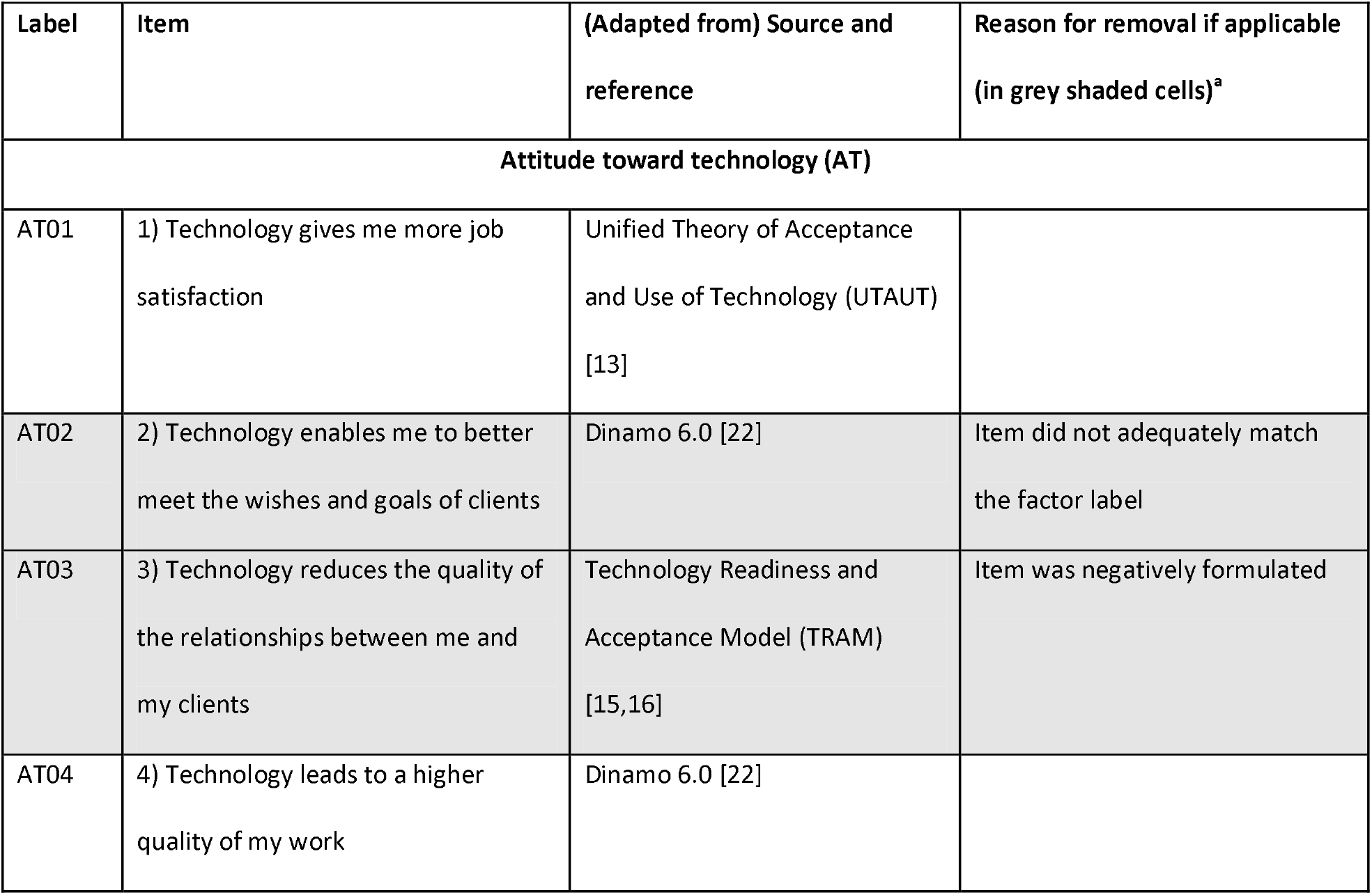
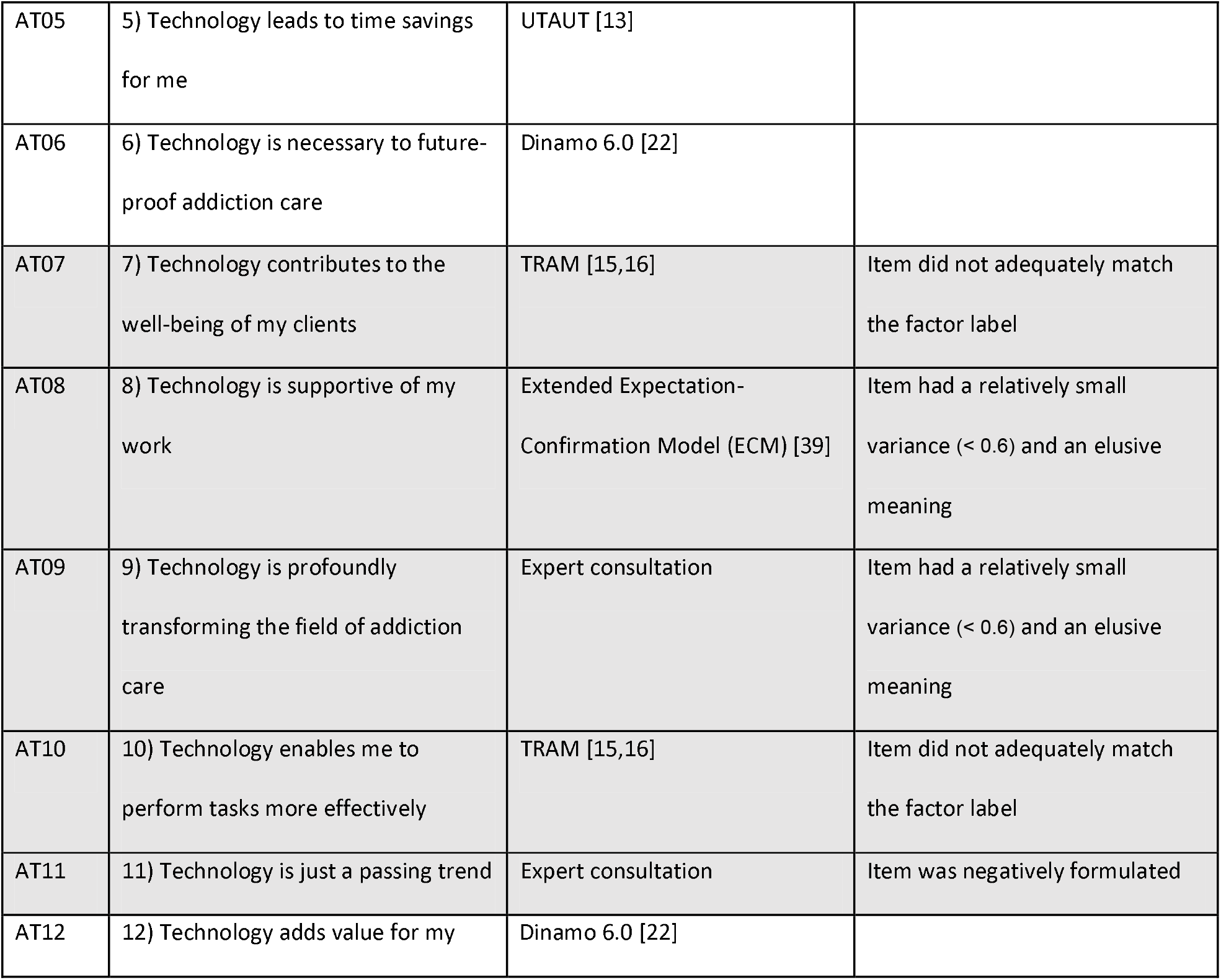
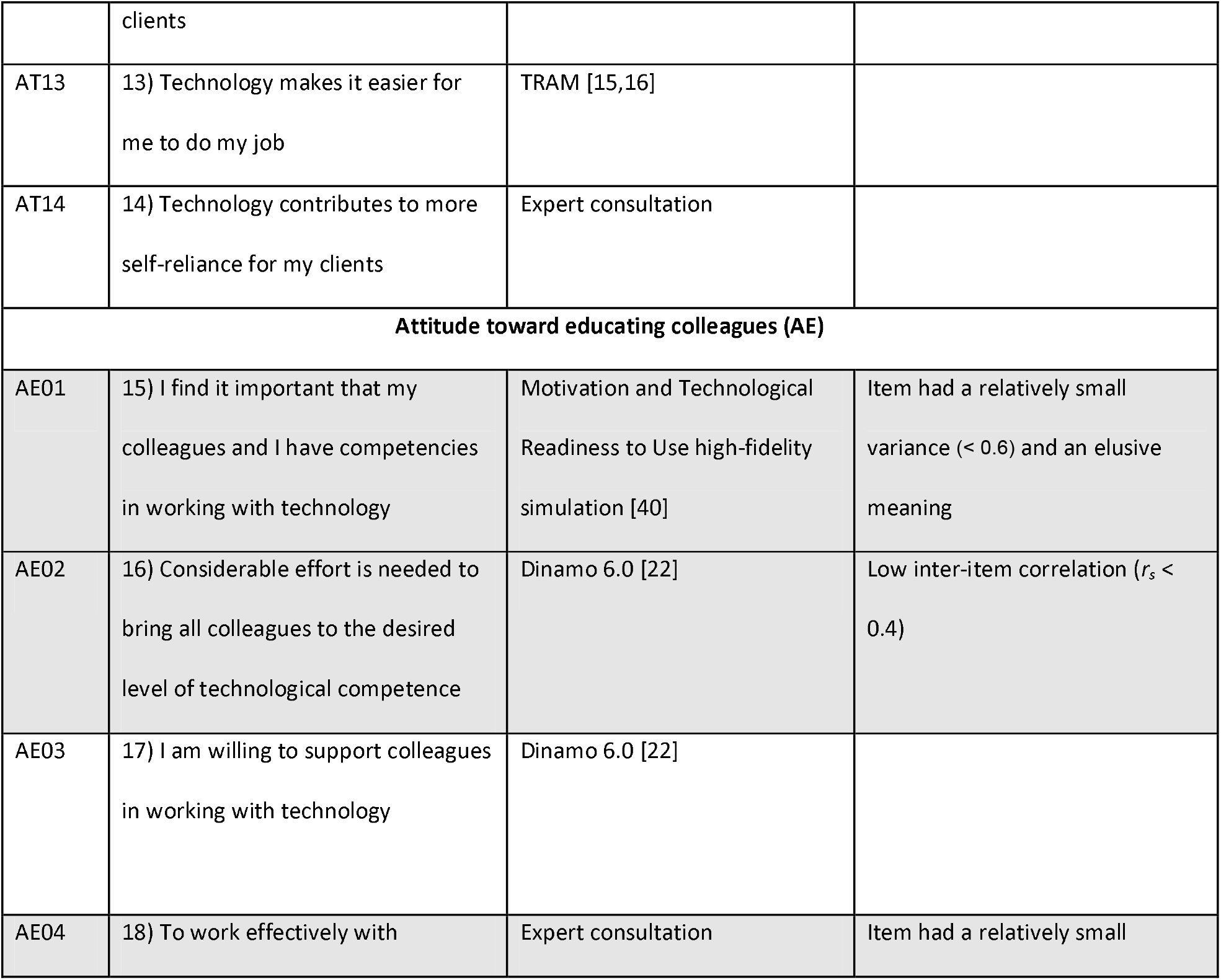
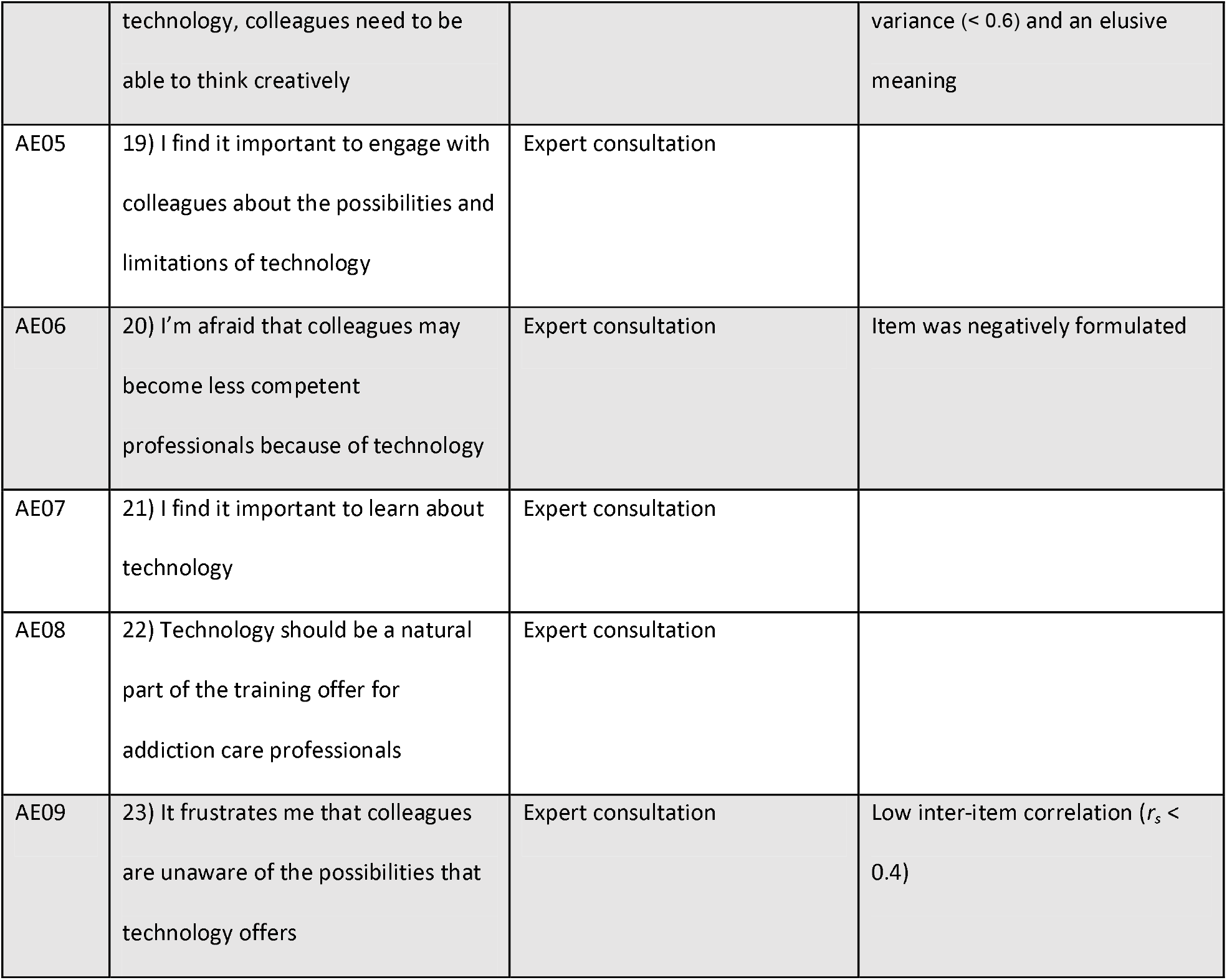
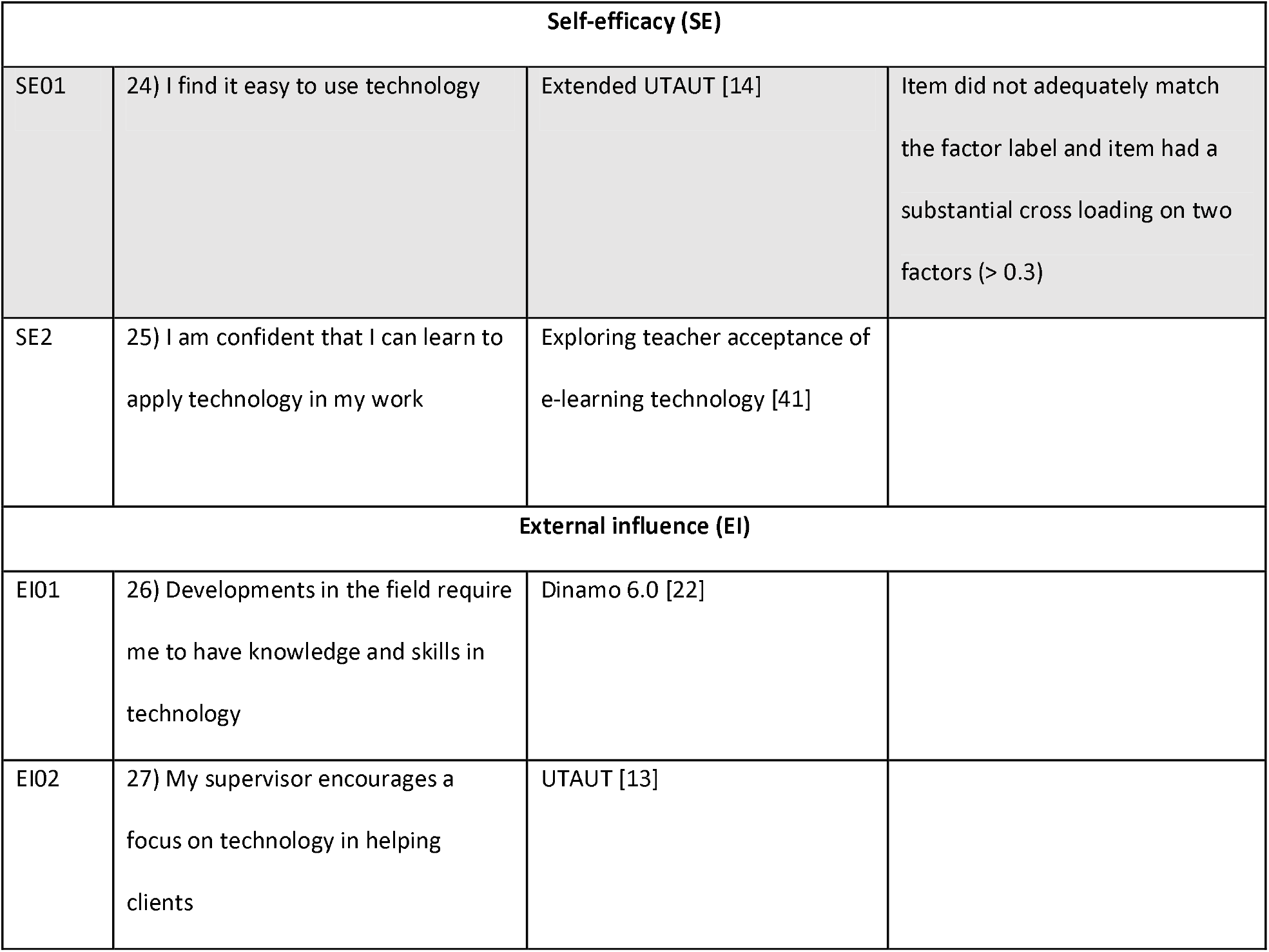
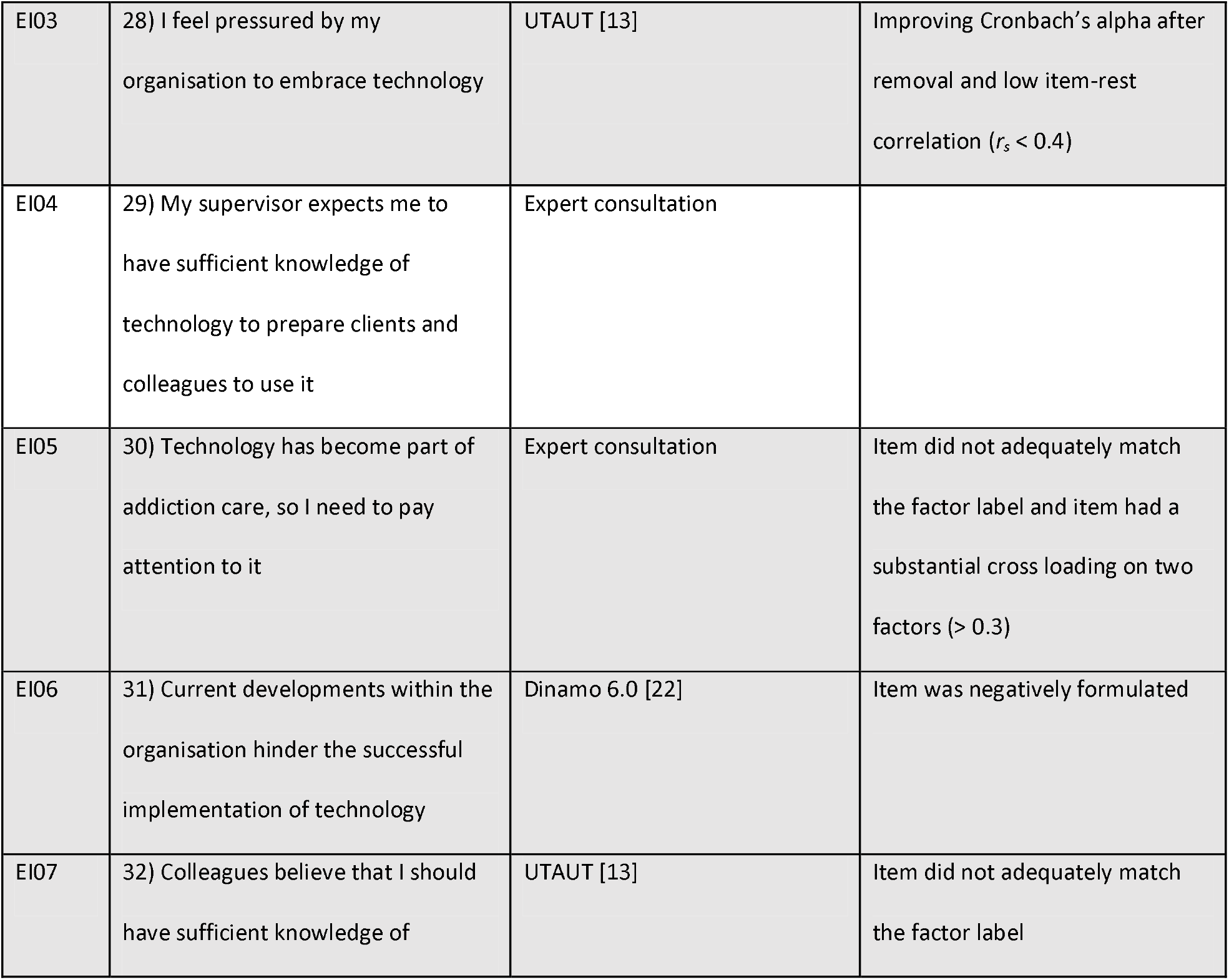
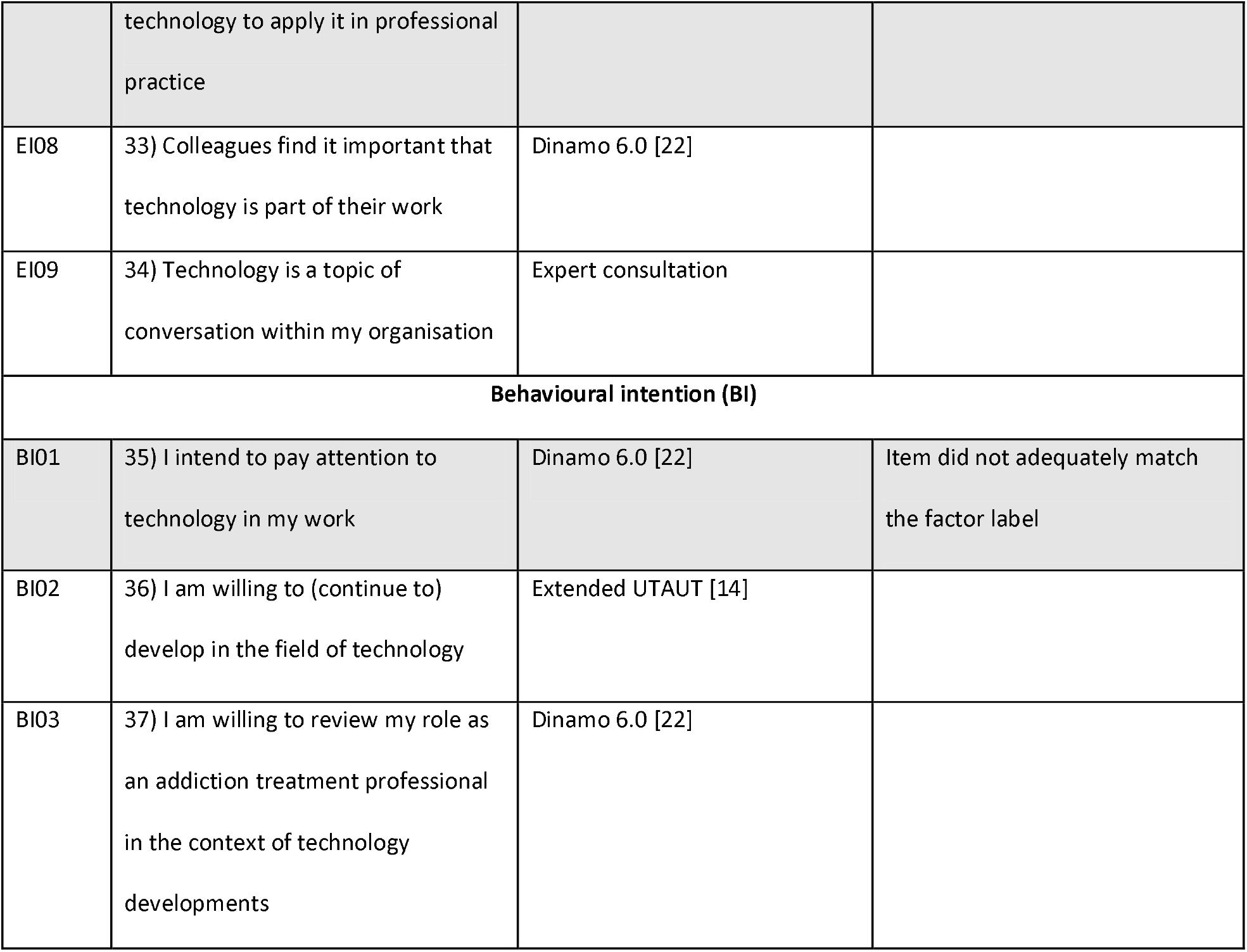
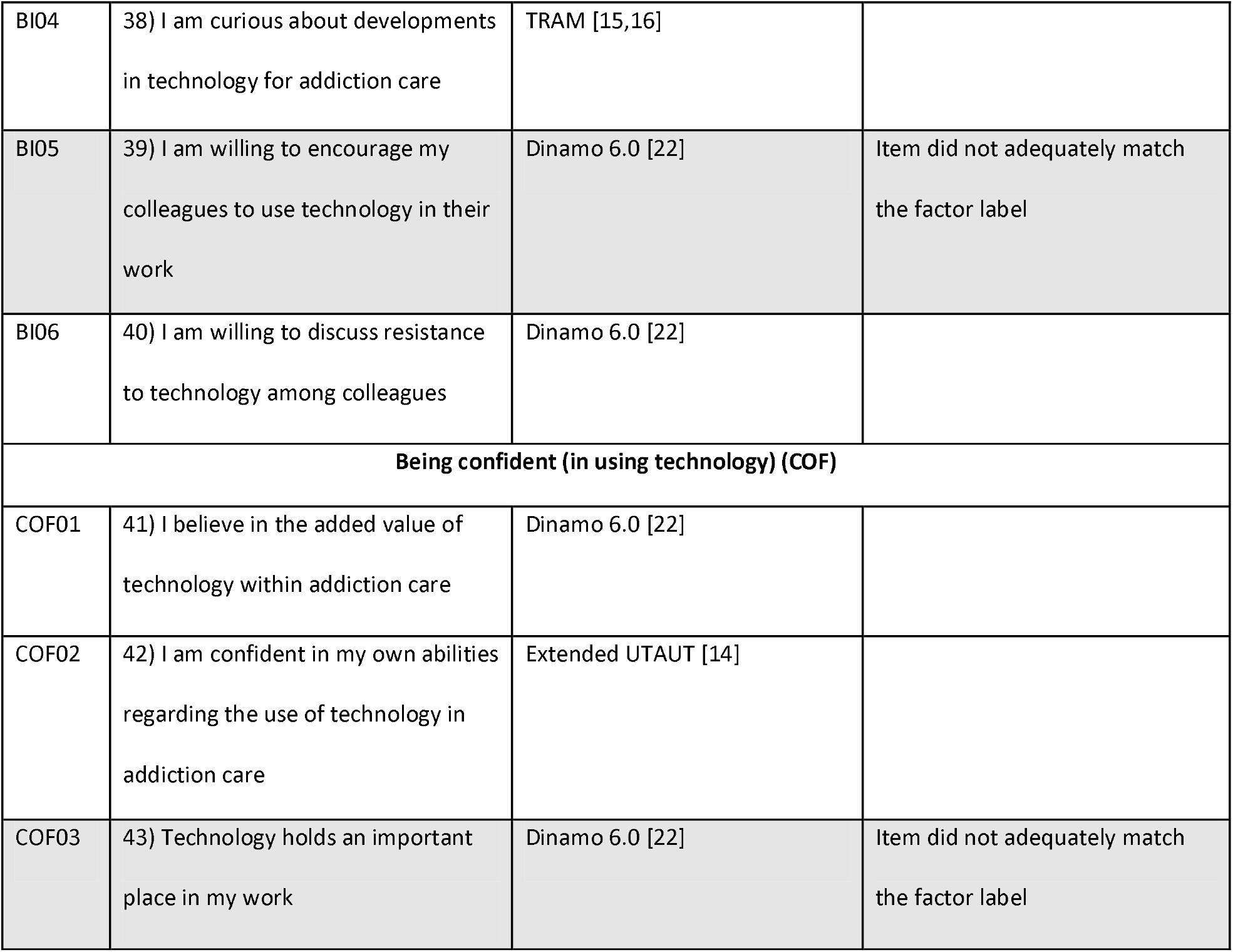
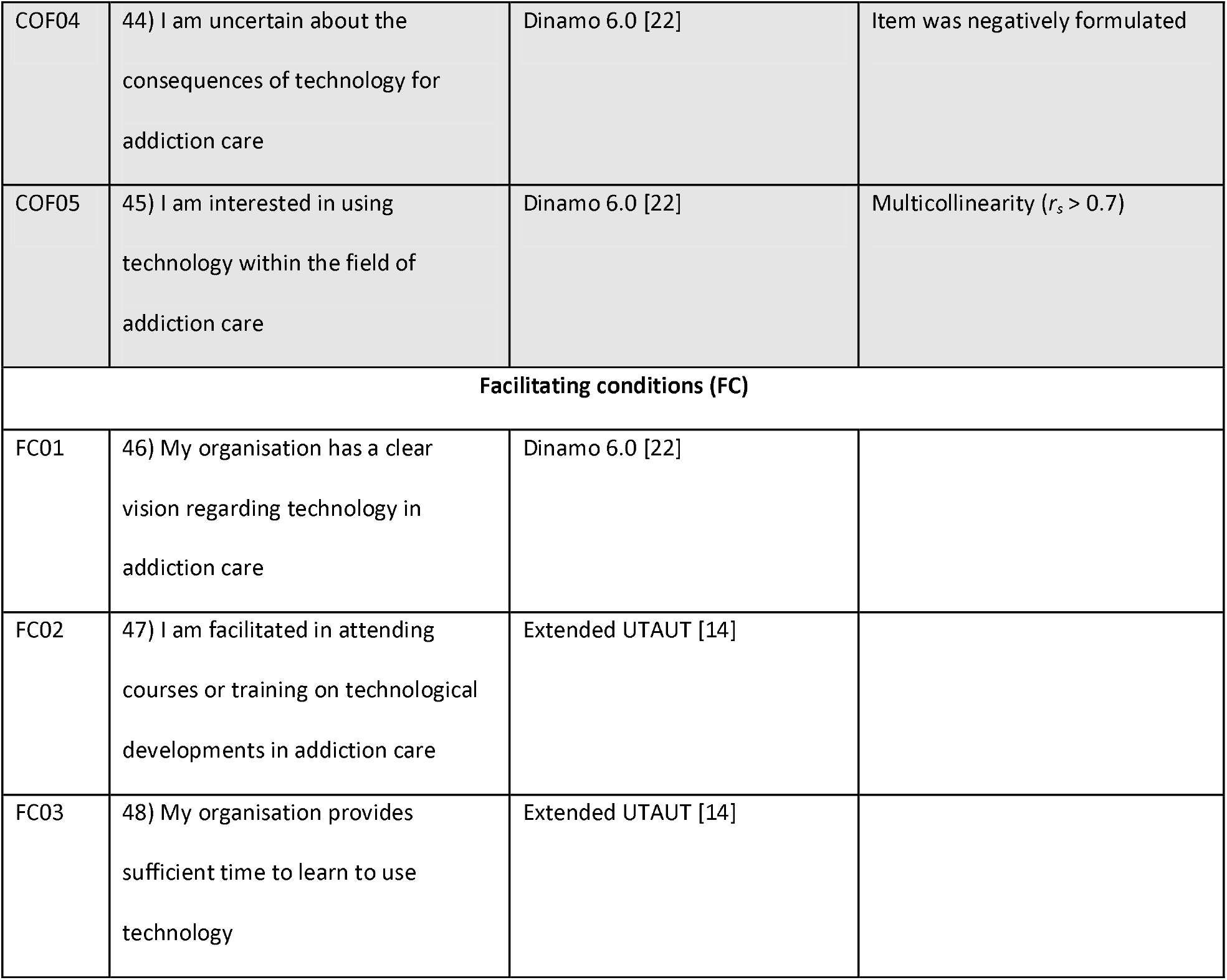
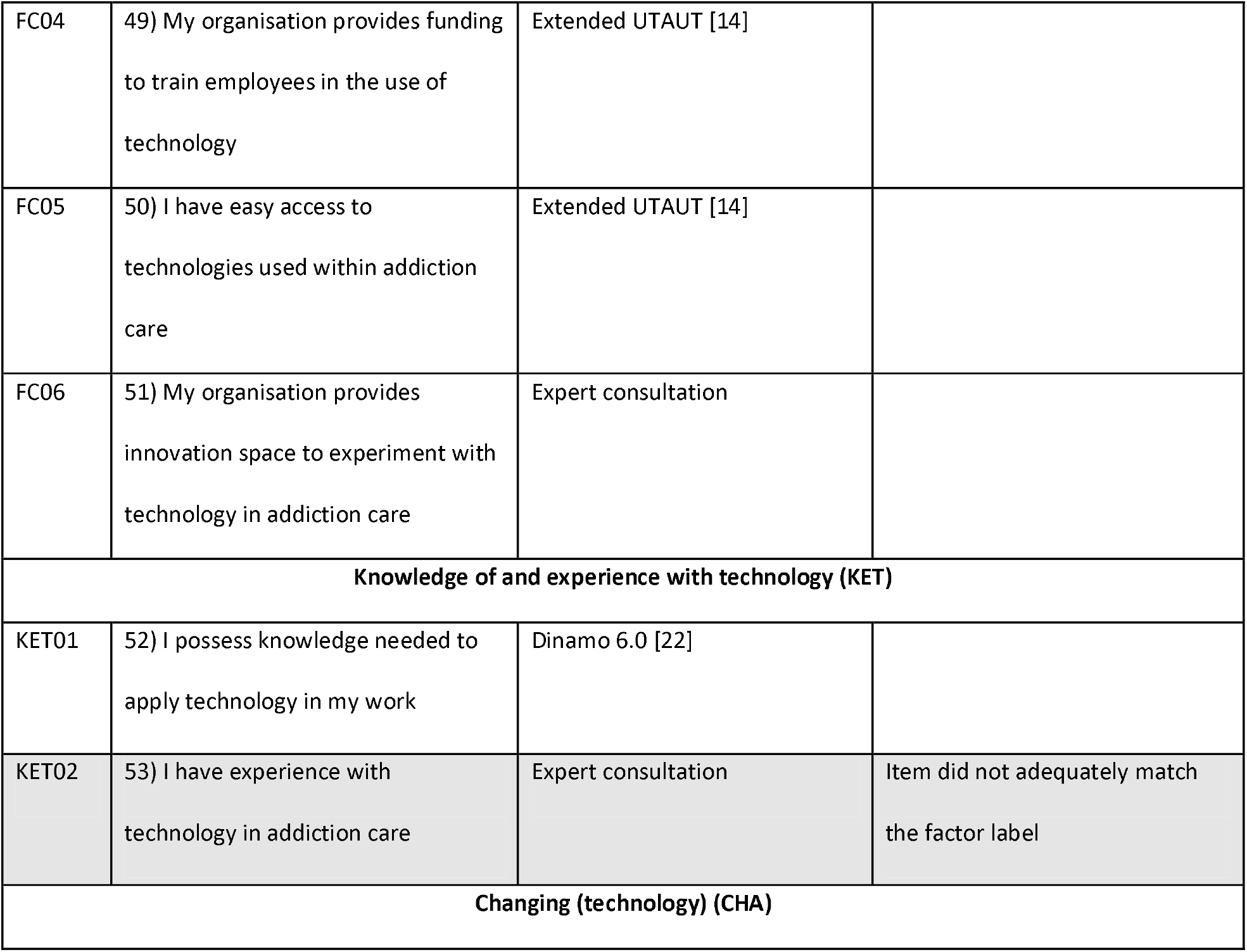
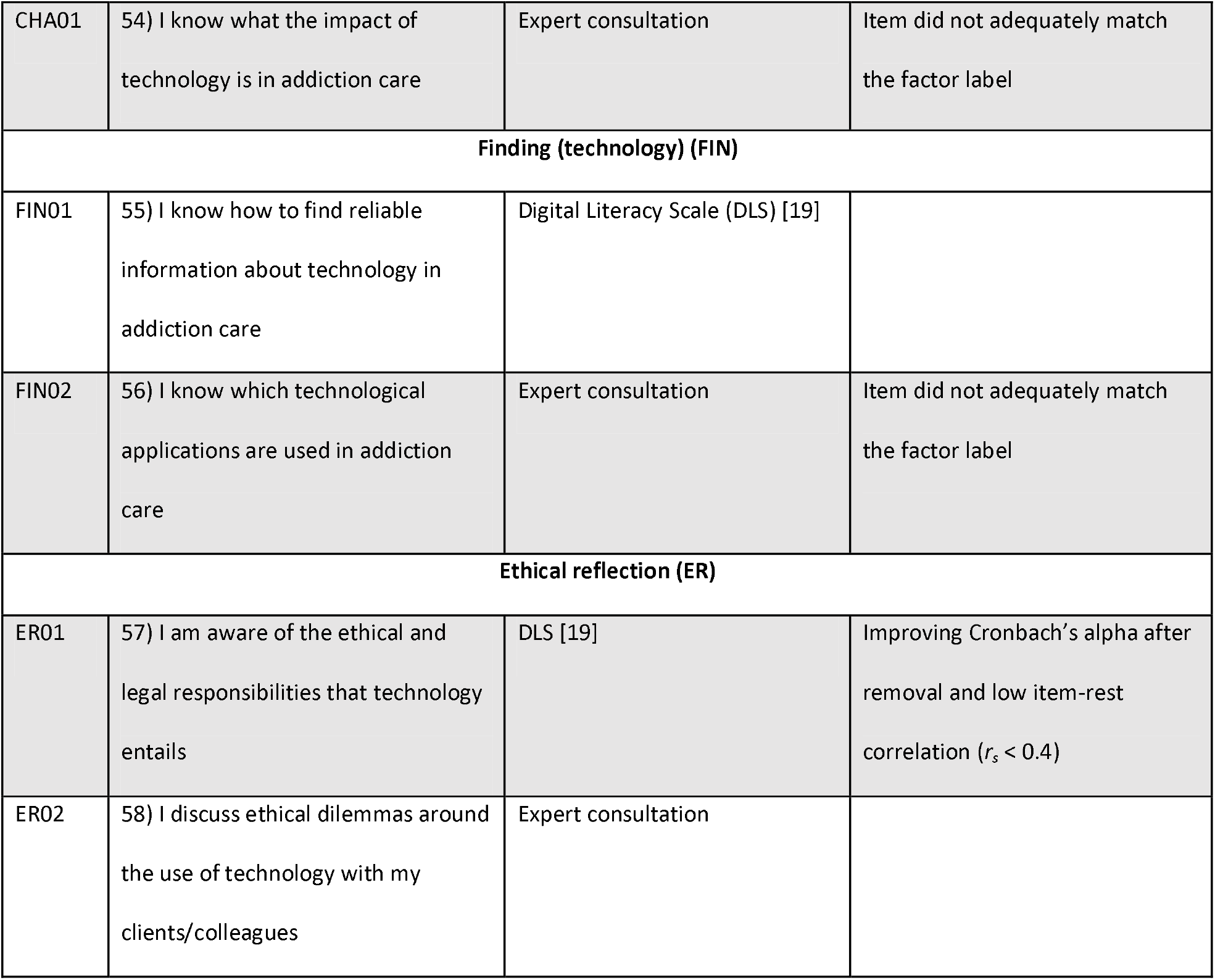
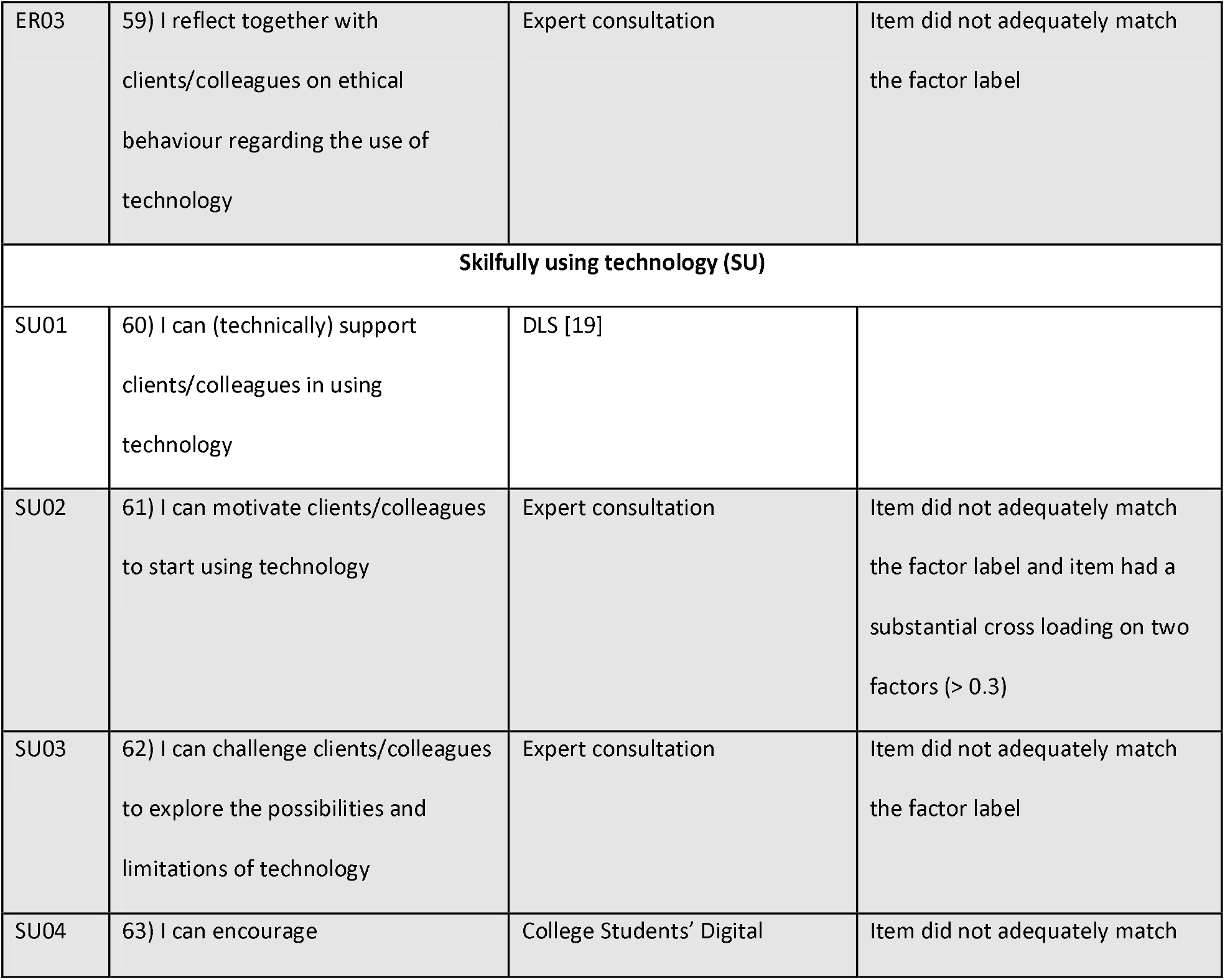
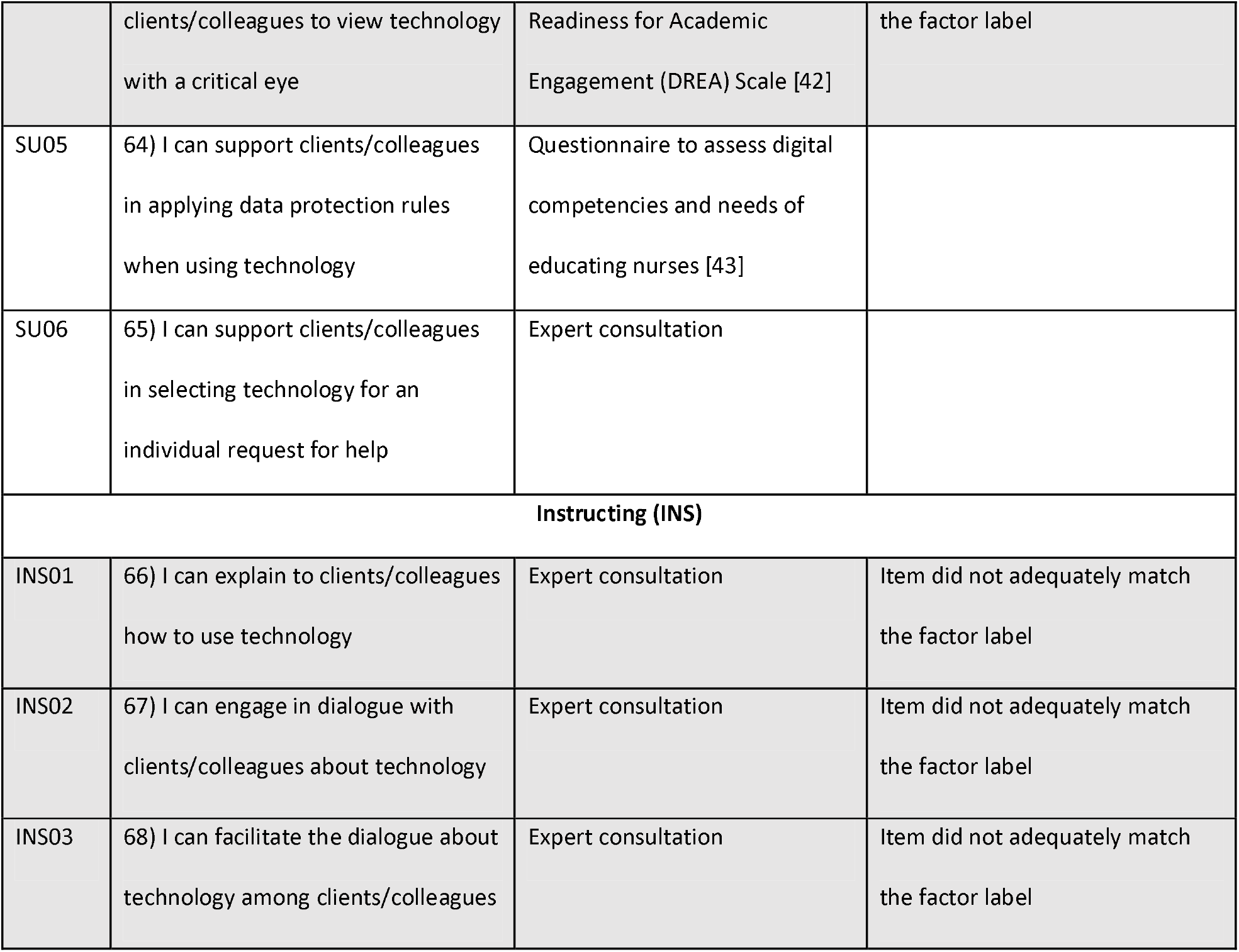
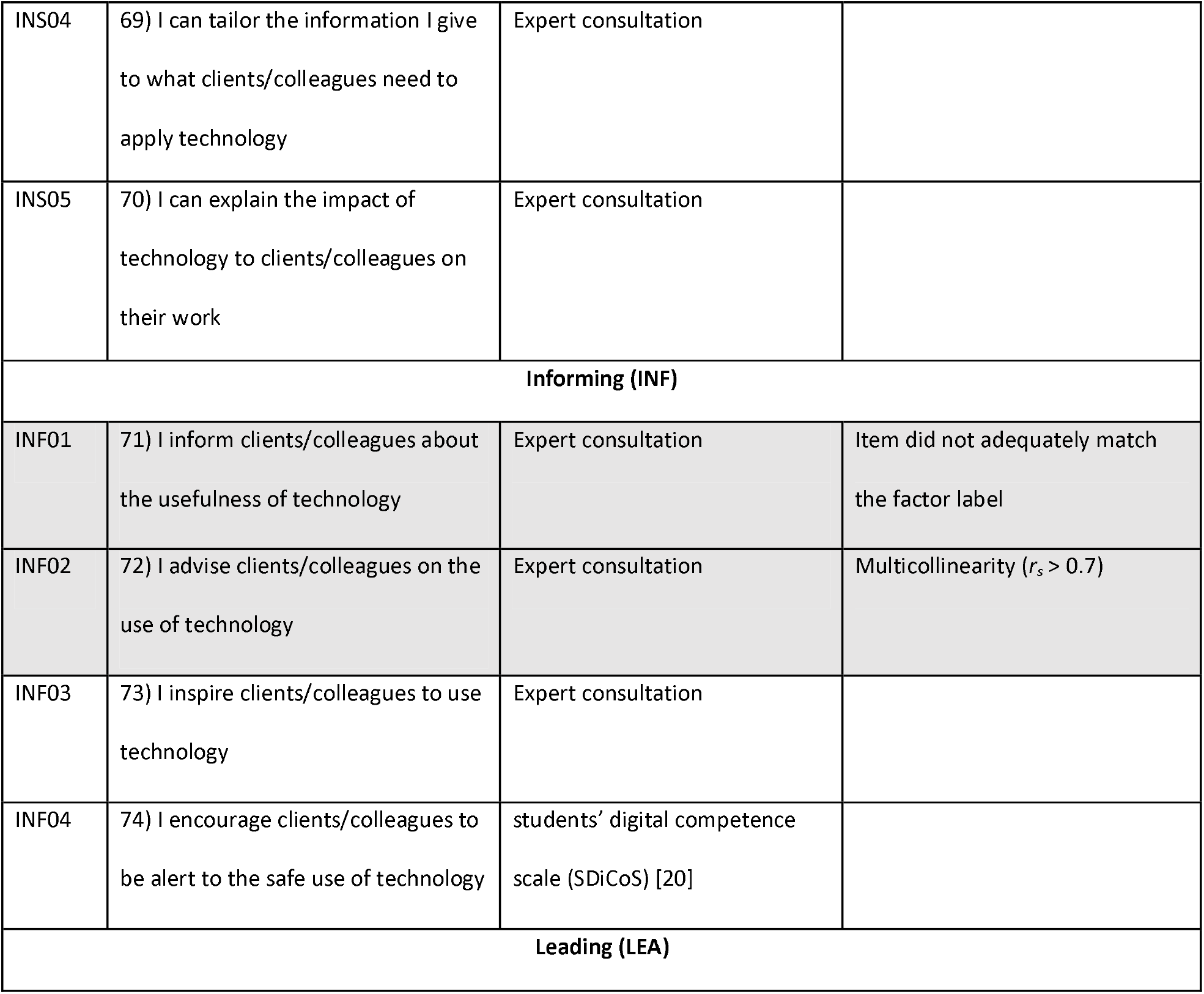
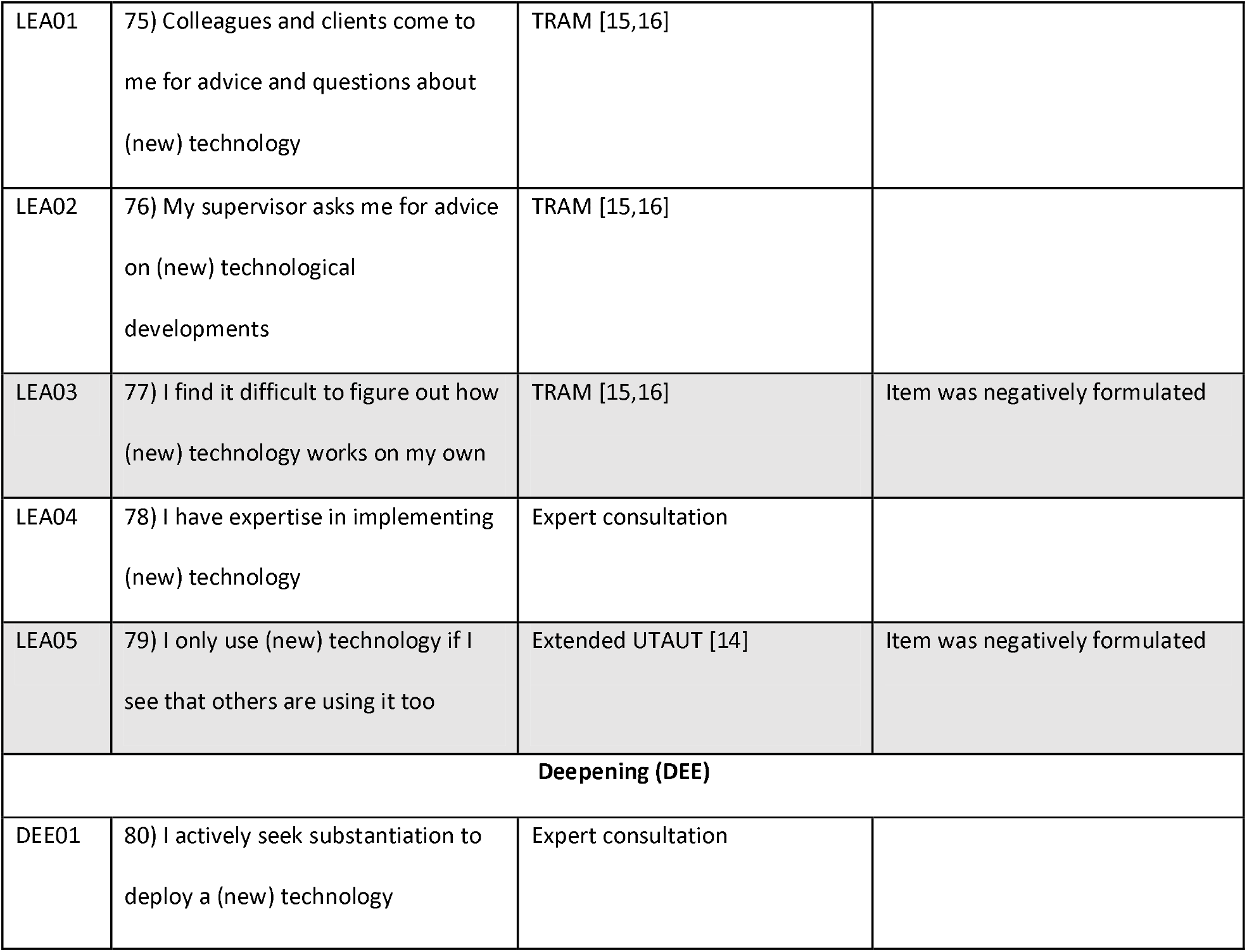
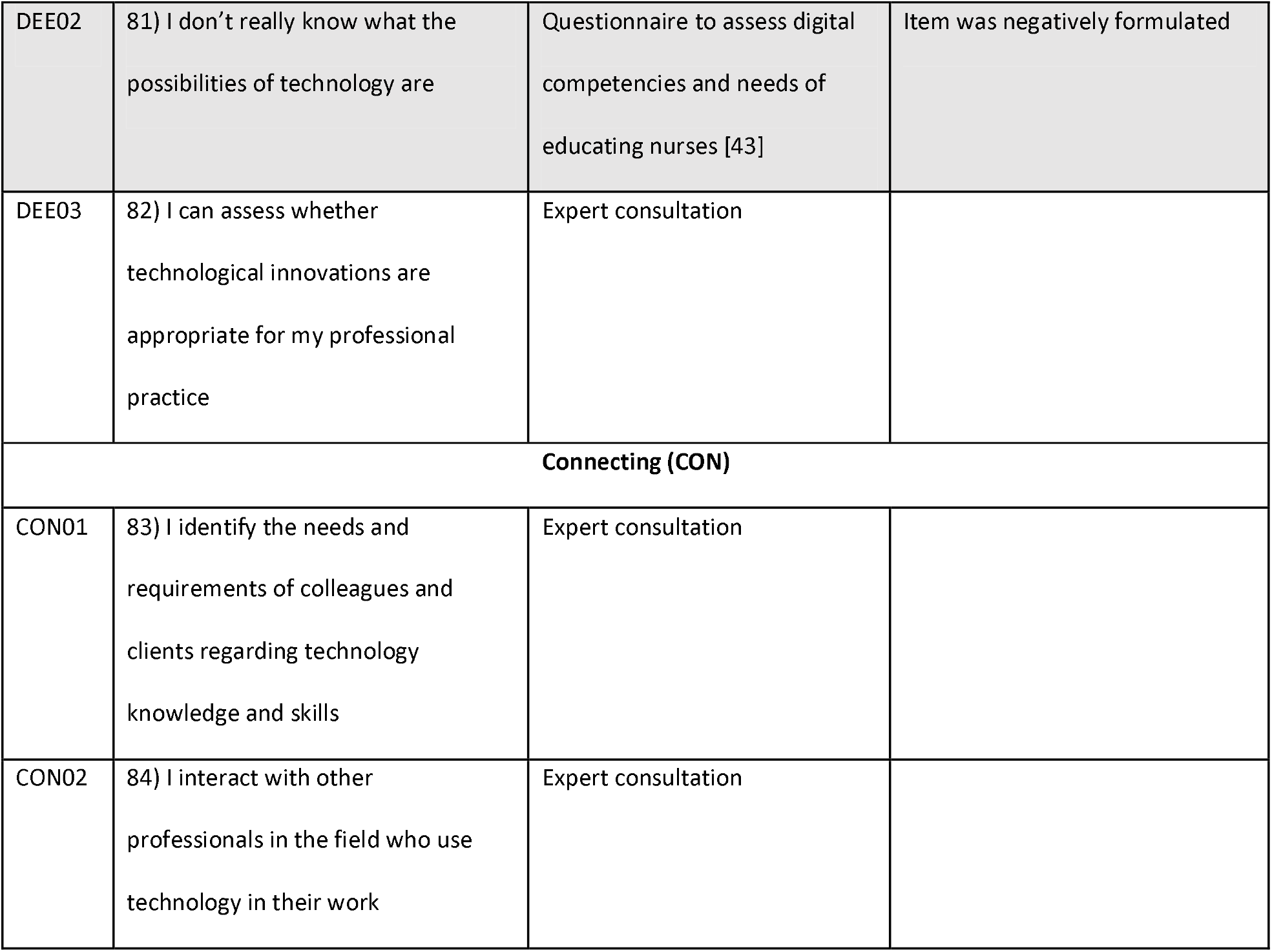
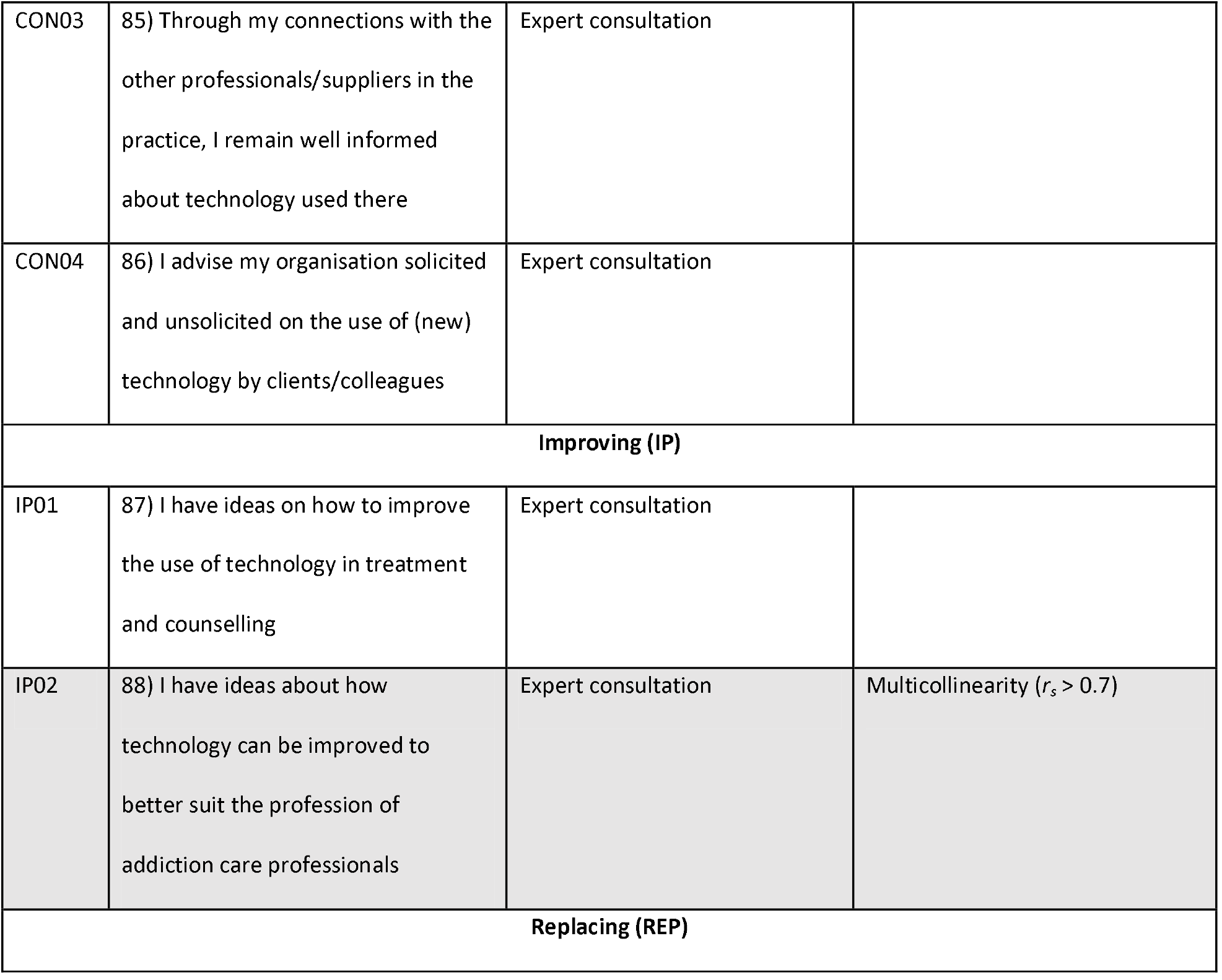
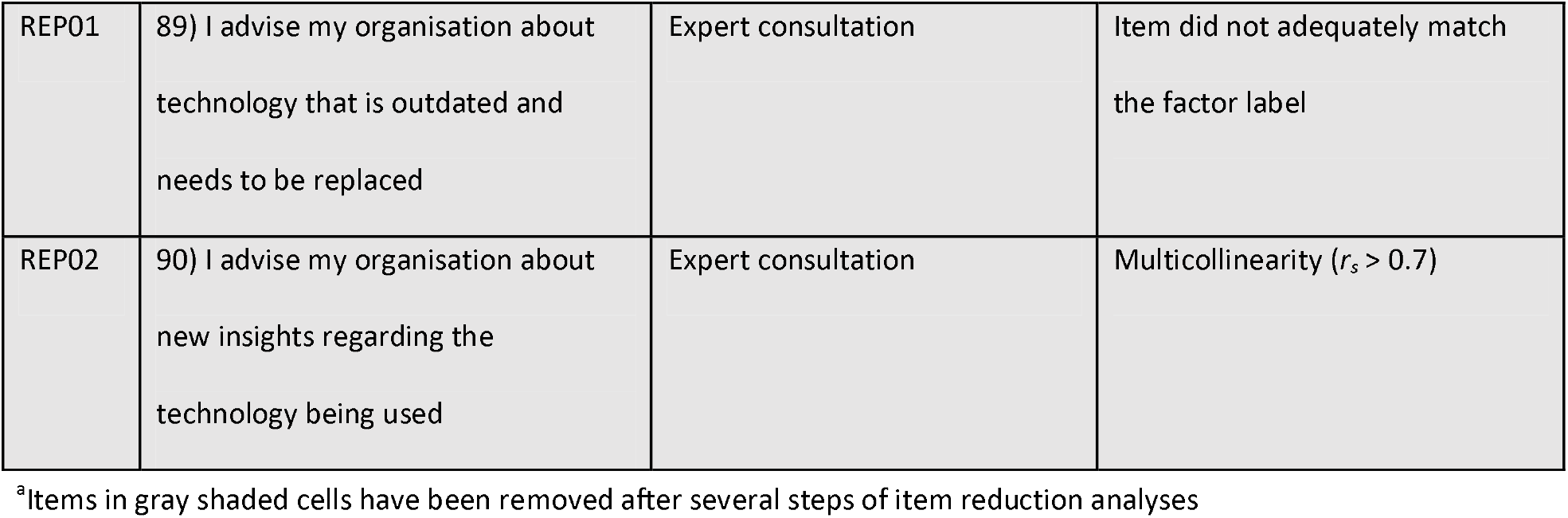
Original item set, their sources and reason for removal if applicable.

**S2 Table.**
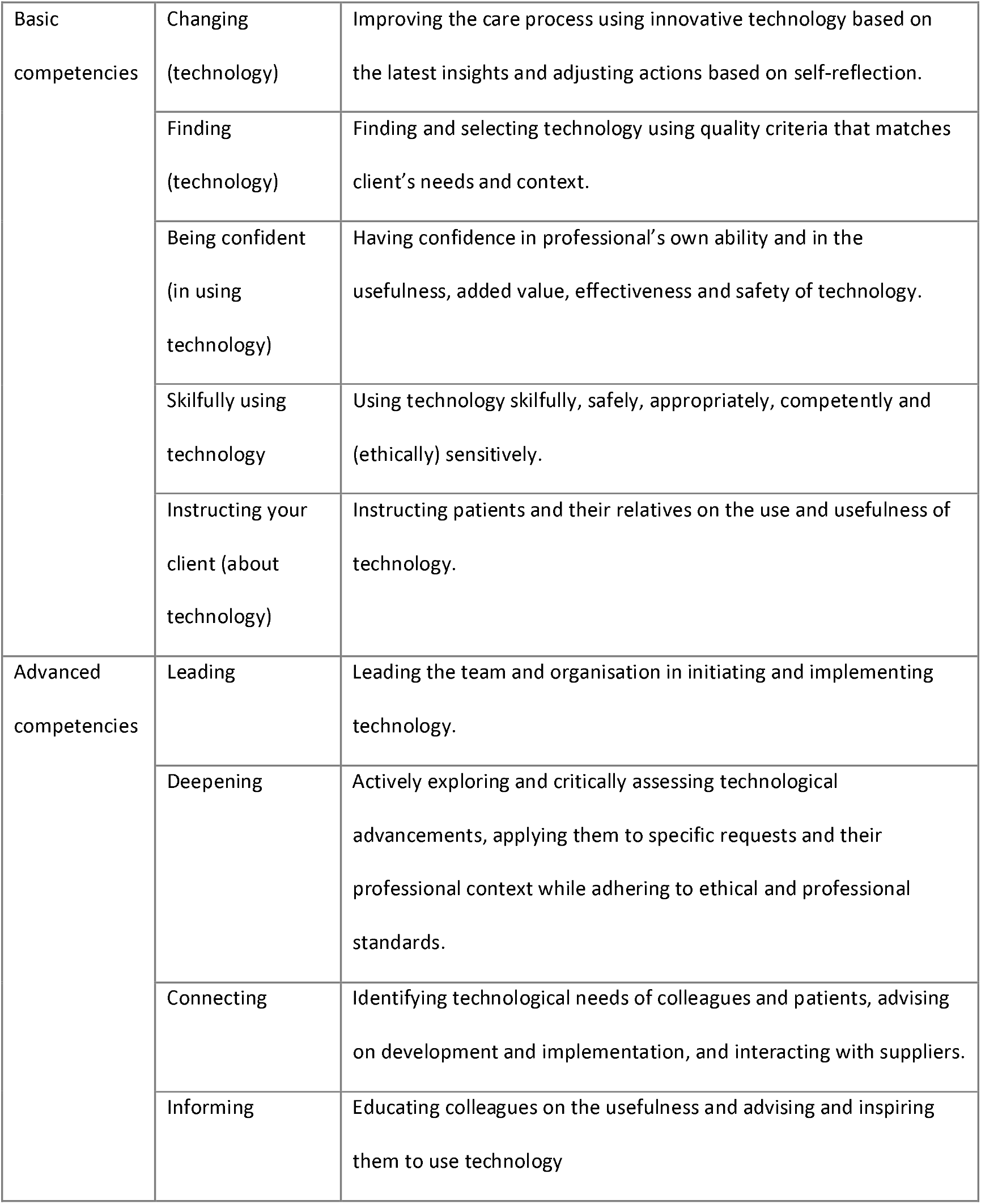
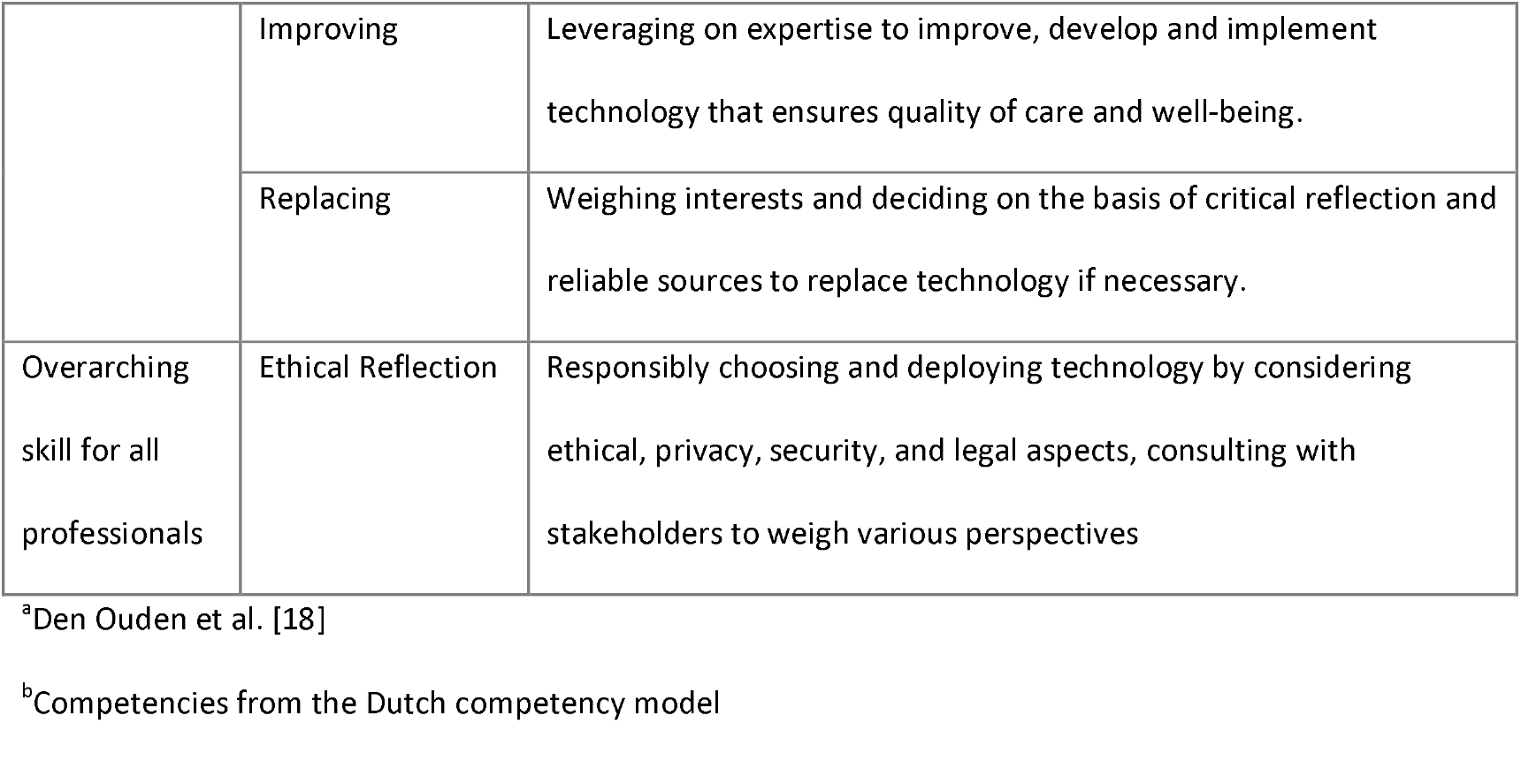
Description of technology competencies of health care professionals ^a^.

## References

1. Keij B, Versluis A, Alblas E, Keuper J, LHD van Tuyl, van der Vaart R. [eHealth monitor 2023. State of affairs in digital healthcare] [Internet]. Rijksinstituut voor Volksgezondheid en Milieu RIVM; 2024 [cited 2025 Apr 9]. Dutch. Available from: https://rivm.openrepository.com/handle/10029/627502

2. Kooij L, Van Harten WH. Healthcare and Digital Transformation. In: Kip H, Beerlage-De Jong N, Van Gemert-Pijnen LJEWC, Sanderman R, Kelders SM, editors. eHealth Research, Theory and Development A Multidisciplinary Approach. Second Edition. Abingdon, Oxon: Routledge; 2024. p. 53–70.

3. Van Gemert-Pijnen JEWC, Kip H, Kelders SM, Sanderman R, Beerlage-de Jong N. Introducing eHealth. In: Kip H, Beerlage-de Jong N, Van Gemert-Pijnen JEWC, Sanderman R, Kelders SM, editors. eHealth Research Theory and Development A Multidisciplinary Approach. 2nd ed. London and New York: Routledge; 2024. p. 3–19.

4. Greenhalgh T, Abimbola S. The NASSS Framework – A synthesis of multiple theories of technology implementation. Stud Health Technol Inform. 2019;263:193–204.

5. Plsek PE, Greenhalgh T. Complexity science: The challenge of complexity in health care. BMJ. 2001 Sept 15;323(7313):625–8.

6. Schreiweis B, Pobiruchin M, Strotbaum V, Suleder J, Wiesner M, Bergh B. Barriers and Facilitators to the Implementation of eHealth Services: Systematic Literature Analysis. J Med Internet Res. 2019 Nov 22;21(11):e14197.

7. Alotaibi N, Wilson CB, Traynor M. Enhancing digital readiness and capability in healthcare: a systematic review of interventions, barriers, and facilitators. BMC Health Serv Res. 2025 Apr 4;25(1):500.

8. Pieterse M, Braakman-Jansen A, Cruz Martinez RR, Van Gemert-Pijnen L (J. EWC). Sustainable eHealth Implementation A Practical Perspective. In: eHealth Research Theory and Development [Internet]. 2nd edn London: Routledge; 2024 [cited 2025 May 14]. p. 237–50. Available from: https://www.taylorfrancis.com/books/9781003302049/chapters/10.4324/9781003302049-16

9. Kelders SM, Perski O. Engagement. In: Kip H, Beerlage-De Jong N, Van Gemert-Pijnen LJEWC, Sanderman R, Kelders SM, editors. eHealth Research, Theory and Development A Multidisciplinary Approach. Second Edition. Abingdon, Oxon: Routledge; 2024. p. 251–68.

10. Cannavacciuolo L, Capaldo G, Ponsiglione C. Digital innovation and organizational changes in the healthcare sector: Multiple case studies of telemedicine project implementation. Technovation. 2023 Feb;120:102550.

11. Postel M, Jaschinski C, Van Til J, Wentzel J, De Vries S, Den Ouden M. [wanting and being able: technology competence scan for educators in healthcare and welbeing]. Onderwijs en gezondheidszorg. 2024;48(2):22–5. Dutch

12. Artino AR, La Rochelle JS, Dezee KJ, Gehlbach H. Developing questionnaires for educational research: AMEE Guide No. 87. Med Teach. 2014 June;36(6):463–74.

13. Venkatesh, Morris, Davis, Davis. User Acceptance of Information Technology: Toward a Unified View. MIS Q. 2003;27(3):425.

14. Venkatesh, Thong, Xu. Consumer Acceptance and Use of Information Technology: Extending the Unified Theory of Acceptance and Use of Technology. MIS Q. 2012;36(1):157.

15. Buyle R, Van Compernolle M, Vlassenroot E, Vanlishout Z, Mechant P, Mannens E. “Technology Readiness and Acceptance Model” as a Predictor for the Use Intention of Data Standards in Smart Cities. Media Commun. 2018 Dec 21;6(4):127–39.

16. Lin C, Shih H, Sher PJ. Integrating technology readiness into technology acceptance: The TRAM model. Psychol Mark. 2007 July;24(7):641–57.

17. Heijblom M, Beijer J, Van Dijk J, Keune M. [Technology in Saxion University of Applied Sciences nursing education]. Deventer/Enschede: Saxion, Academie Gezondheidszorg; 2015. Dutch

18. Den Ouden MEM, Stokkers-Scholten J, Jaschinski C, Oosterbroek AR, Groeneveld S, Van Os-Medendorp H, et al. [V-model: Technology competencies of healthcare and wellbeing professionals of today and tomorrow]. Onderwijs En Gezondheidszorg. 2023;47(4):22–5. Dutch

19. Bayrakcı S, Narmanlıoğlu H. Digital Literacy as Whole of Digital Competences: Scale Development Study. Journal of thought and society. 2021 June 29;3:1–30.

20. Tzafilkou K, Perifanou M, Economides AA. Development and validation of students’ digital competence scale (SDiCoS). Int J Educ Technol High Educ. 2022 Dec;19(1):30.

21. Tactus 125 jaar [Internet]. Tactus Verslavingszorg [Tactus Addiction Care]. [cited 2025 Aug 29]. Dutch. Available from: https://www.tactus.nl/tactus-125-jaar/

22. Change Manager BV. In Metselaar (1997). Dinamo 6.0 [Internet]. Change Manager BV; 2017. Available from: http://hoadd.noordhoff.nl/sites/7606/_assets/7606d16.pdf

23. IBM Corp. IBM SPSS Statistics for Windows, Version 29.0 [Internet]. Armonk, NY: IBM Corp; 2022. Available from: https://www.ibm.com/products/spss-statistics

24. Artino AR, Gehlbach H, Durning SJ. AM last page: Avoiding five common pitfalls of survey design. Acad Med J Assoc Am Med Coll. 2011 Oct;86(10):1327.

25. DiStefano C, Motl RW. Further Investigating Method Effects Associated With Negatively Worded Items on Self-Report Surveys. Struct Equ Model Multidiscip J. 2006 June 28;13(3):440– 64.

26. Marjanovic Z, Maidens AL. A Clarified Examination of the Item Wording Effect: Item Valence (Good vs. Bad) Versus Semantic Framing (I Am vs. I Am Not). Psychol Rep. 2024 Nov 14;00332941241301353.

27. Kaiser HF, Rice J. Little Jiffy, Mark Iv. Educ Psychol Meas. 1974 Apr;34(1):111–7.

28. Cattell RB. The Scree Test For The Number Of Factors. Multivar Behav Res. 1966 Apr;1(2):245– 76.

29. Terwee CB, Bot SDM, De Boer MR, Van Der Windt DAWM, Knol DL, Dekker J, et al. Quality criteria were proposed for measurement properties of health status questionnaires. J Clin Epidemiol. 2007 Jan;60(1):34–42.

30. Kyriazos TA, Stalikas A. Applied Psychometrics: The Steps of Scale Development and Standardization Process. Psychology. 2018;09(11):2531–60.

31. Ajzen I. The theory of planned behavior. Organ Behav Hum Decis Process. 1991 Dec;50(2):179– 211.

32. Davis FD. Perceived Usefulness, Perceived Ease of Use, and User Acceptance of Information Technology. MIS Q. 1989 Sept;13(3):319.

33. Van Der Vaart R, Atema V, Evers AWM. Guided online self-management interventions in primary care: a survey on use, facilitators, and barriers. BMC Fam Pract. 2016 Dec;17(1):27.

34. Boateng GO, Neilands TB, Frongillo EA, Melgar-Quiñonez HR, Young SL. Best Practices for Developing and Validating Scales for Health, Social, and Behavioral Research: A Primer. Front Public Health. 2018 June 11;6:149.

35. Bryman A. Social research methods. Fifth edition. Oxford: Oxford University Press; 2016. 747 p.

36. Nunnally JC. Psychometric theory. 2. ed. New York: McGraw-Hill; 1978. 701 p. (McGraw-Hill series in psychology).

37. Field A. Discovering statistics using IBM SPSS statistics. 5th edition. Los Angeles London New Delhi Singapore Washington DC Melbourne: SAGE; 2018. 1070 p. (SAGE edge).

38. Comrey AL, Lee HB. A First Course in Factor Analysis. 2nd ed. Lawrence Erlbaum Associates, Inc.; 1992.

39. Cheng YM. What Drives Nurses’ Blended e-Learning Continuance Intention? J Educ Technol Soc. 2014;14(4):203–15.

40. Duvall JJ. Motivation and technological readiness in the use of high-fidelity simulation: a descriptive comparative study of nurse educators [Internet]. [Tuscaloosa, Alabama]: University of Alabama; 2012 [cited 2025 Apr 11]. Available from: https://irapi.ua.edu/api/core/bitstreams/afdaa75f-68db-4560-a996-0b73211d25b8/content

41. Yuen AHK, Ma WWK. Exploring teacher acceptance of e-learning technology. Asia-Pac J Teach Educ. 2008 Aug;36(3):229–43.

42. Hong AJ, Kim HJ. College Students’ Digital Readiness for Academic Engagement (DRAE) Scale: Scale Development and Validation. Asia-Pac Educ Res. 2018 Aug 1;27(4):303–12.

43. Jobst S, Lindwedel U, Marx H, Pazouki R, Ziegler S, König P, et al. Competencies and needs of nurse educators and clinical mentors for teaching in the digital age – a multi-institutional, cross-sectional study. BMC Nurs. 2022 Aug 28;21(1):1–13.

