## Supplemental Table 1 for "Development of the Technology Readiness Inventory for Professionals (TRIP): field testing in Dutch addiction care professionals"

### S1 Table. Original item set, their sources and reason for removal if applicable

| Label | Item | (Adapted from) Source and reference | Reason for removal if applicable (in grey shaded cells)[a] |
|---|---|---|---|
| | | **Attitude toward technology (AT)** | |
| AT01 | 1) Technology gives me more job satisfaction | Unified Theory of Acceptance and Use of Technology (UTAUT) [13] | |
| AT02 | 2) Technology enables me to better meet the wishes and goals of clients | Dinamo 6.0 [22] | Item did not adequately match the factor label |
| AT03 | 3) Technology reduces the quality of the relationships between me and my clients | Technology Readiness and Acceptance Model (TRAM) [15,16] | Item was negatively formulated |
| AT04 | 4) Technology leads to a higher quality of my work | Dinamo 6.0 [22] | |

| | | | |
|---|---|---|---|
| AT05 | 5) Technology leads to time savings for me | UTAUT [13] | |
| AT06 | 6) Technology is necessary to future-proof addiction care | Dinamo 6.0 [22] | |
| AT07 | 7) Technology contributes to the well-being of my clients | TRAM [15,16] | Item did not adequately match the factor label |
| AT08 | 8) Technology is supportive of my work | Extended Expectation-Confirmation Model (ECM) [39] | Item had a relatively small variance (< 0.6) and an elusive meaning |
| AT09 | 9) Technology is profoundly transforming the field of addiction care | Expert consultation | Item had a relatively small variance (< 0.6) and an elusive meaning |
| AT10 | 10) Technology enables me to perform tasks more effectively | TRAM [15,16] | Item did not adequately match the factor label |
| AT11 | 11) Technology is just a passing trend | Expert consultation | Item was negatively formulated |

| | | | |
|---|---|---|---|
| AT12 | 12) Technology adds value for my clients | Dinamo 6.0 [22] | |
| AT13 | 13) Technology makes it easier for me to do my job | TRAM [15,16] | |
| AT14 | 14) Technology contributes to more self-reliance for my clients | Expert consultation | |
| **Attitude toward educating colleagues (AE)** | | | |
| AE01 | 15) I find it important that my colleagues and I have competencies in working with technology | Motivation and Technological Readiness to Use high-fidelity simulation [40] | Item had a relatively small variance (< 0.6) and an elusive meaning |
| AE02 | 16) Considerable effort is needed to bring all colleagues to the desired level of technological competence | Dinamo 6.0 [22] | Low inter-item correlation ($r_s$ < 0.4) |
| AE03 | 17) I am willing to support colleagues in working with technology | Dinamo 6.0 [22] | |

| | | | |
|---|---|---|---|
| AE04 | 18) To work effectively with technology, colleagues need to be able to think creatively | Expert consultation | Item had a relatively small variance (< 0.6) and an elusive meaning |
| AE05 | 19) I find it important to engage with colleagues about the possibilities and limitations of technology | Expert consultation | |
| AE06 | 20) I'm afraid that colleagues may become less competent professionals because of technology | Expert consultation | Item was negatively formulated |
| AE07 | 21) I find it important to learn about technology | Expert consultation | |
| AE08 | 22) Technology should be a natural part of the training offer for addiction care professionals | Expert consultation | |

| | | | |
|---|---|---|---|
| AE09 | 23) It frustrates me that colleagues are unaware of the possibilities that technology offers | Expert consultation | Low inter-item correlation ($r_s$ < 0.4) |
| | **Self-efficacy (SE)** | | |
| SE01 | 24) I find it easy to use technology | Extended UTAUT [14] | Item did not adequately match the factor label and item had a substantial cross loading on two factors (> 0.3) |
| SE2 | 25) I am confident that I can learn to apply technology in my work | Exploring teacher acceptance of e-learning technology [41] | |
| | **External influence (EI)** | | |
| EI01 | 26) Developments in the field require me to have knowledge and skills in technology | Dinamo 6.0 [22] | |

| | | | |
|---|---|---|---|
| EI02 | 27) My supervisor encourages a focus on technology in helping clients | UTAUT [13] | |
| EI03 | 28) I feel pressured by my organisation to embrace technology | UTAUT [13] | Improving Cronbach's alpha after removal and low item-rest correlation ($r_s < 0.4$) |
| EI04 | 29) My supervisor expects me to have sufficient knowledge of technology to prepare clients and colleagues to use it | Expert consultation | |
| EI05 | 30) Technology has become part of addiction care, so I need to pay attention to it | Expert consultation | Item did not adequately match the factor label and item had a substantial cross loading on two factors (> 0.3) |

| | | | |
|---|---|---|---|
| EI06 | 31) Current developments within the organisation hinder the successful implementation of technology | Dinamo 6.0 [22] | Item was negatively formulated |
| EI07 | 32) Colleagues believe that I should have sufficient knowledge of technology to apply it in professional practice | UTAUT [13] | Item did not adequately match the factor label |
| EI08 | 33) Colleagues find it important that technology is part of their work | Dinamo 6.0 [22] | |
| EI09 | 34) Technology is a topic of conversation within my organisation | Expert consultation | |
| **Behavioural intention (BI)** | | | |
| BI01 | 35) I intend to pay attention to technology in my work | Dinamo 6.0 [22] | Item did not adequately match the factor label |
| BI02 | 36) I am willing to (continue to) develop in the field of technology | Extended UTAUT [14] | |

| | | | |
|---|---|---|---|
| BI03 | 37) I am willing to review my role as an addiction treatment professional in the context of technology developments | Dinamo 6.0 [22] | |
| BI04 | 38) I am curious about developments in technology for addiction care | TRAM [15,16] | |
| BI05 | 39) I am willing to encourage my colleagues to use technology in their work | Dinamo 6.0 [22] | Item did not adequately match the factor label |
| BI06 | 40) I am willing to discuss resistance to technology among colleagues | Dinamo 6.0 [22] | |
| **Being confident (in using technology) (COF)** | | | |
| COF01 | 41) I believe in the added value of technology within addiction care | Dinamo 6.0 [22] | |

| | | | |
|---|---|---|---|
| COF02 | 42) I am confident in my own abilities regarding the use of technology in addiction care | Extended UTAUT [14] | |
| COF03 | 43) Technology holds an important place in my work | Dinamo 6.0 [22] | Item did not adequately match the factor label |
| COF04 | 44) I am uncertain about the consequences of technology for addiction care | Dinamo 6.0 [22] | Item was negatively formulated |
| COF05 | 45) I am interested in using technology within the field of addiction care | Dinamo 6.0 [22] | Multicollinearity ($r_s > 0.7$) |
| **Facilitating conditions (FC)** | | | |
| FC01 | 46) My organisation has a clear vision regarding technology in addiction care | Dinamo 6.0 [22] | |

| | | |
|---|---|---|
| FC02 | 47) I am facilitated in attending courses or training on technological developments in addiction care | Extended UTAUT [14] |
| FC03 | 48) My organisation provides sufficient time to learn to use technology | Extended UTAUT [14] |
| FC04 | 49) My organisation provides funding to train employees in the use of technology | Extended UTAUT [14] |
| FC05 | 50) I have easy access to technologies used within addiction care | Extended UTAUT [14] |
| FC06 | 51) My organisation provides innovation space to experiment with technology in addiction care | Expert consultation |
| **Knowledge of and experience with technology (KET)** | | |

| | | | |
|---|---|---|---|
| KET01 | 52) I possess knowledge needed to apply technology in my work | Dinamo 6.0 [22] | |
| KET02 | 53) I have experience with technology in addiction care | Expert consultation | Item did not adequately match the factor label |
| **Changing (technology) (CHA)** | | | |
| CHA01 | 54) I know what the impact of technology is in addiction care | Expert consultation | Item did not adequately match the factor label |
| **Finding (technology) (FIN)** | | | |
| FIN01 | 55) I know how to find reliable information about technology in addiction care | Digital Literacy Scale (DLS) [19] | |
| FIN02 | 56) I know which technological applications are used in addiction care | Expert consultation | Item did not adequately match the factor label |
| **Ethical reflection (ER)** | | | |

| | | | |
|---|---|---|---|
| ER01 | 57) I am aware of the ethical and legal responsibilities that technology entails | DLS [19] | Improving Cronbach's alpha after removal and low item-rest correlation ($r_s < 0.4$) |
| ER02 | 58) I discuss ethical dilemmas around the use of technology with my clients/colleagues | Expert consultation | |
| ER03 | 59) I reflect together with clients/colleagues on ethical behaviour regarding the use of technology | Expert consultation | Item did not adequately match the factor label |
| **Skilfully using technology (SU)** | | | |
| SU01 | 60) I can (technically) support clients/colleagues in using technology | DLS [19] | |
| SU02 | 61) I can motivate clients/colleagues to start using technology | Expert consultation | Item did not adequately match the factor label and item had a |

| | | | |
|---|---|---|---|
| | | | substantial cross loading on two factors (> 0.3) |
| SU03 | 62) I can challenge clients/colleagues to explore the possibilities and limitations of technology | Expert consultation | Item did not adequately match the factor label |
| SU04 | 63) I can encourage clients/colleagues to view technology with a critical eye | College Students' Digital Readiness for Academic Engagement (DREA) Scale [42] | Item did not adequately match the factor label |
| SU05 | 64) I can support clients/colleagues in applying data protection rules when using technology | Questionnaire to assess digital competencies and needs of educating nurses [43] | |
| SU06 | 65) I can support clients/colleagues in selecting technology for an individual request for help | Expert consultation | |
| **Instructing (INS)** | | | |

| | | | |
|---|---|---|---|
| INS01 | 66) I can explain to clients/colleagues how to use technology | Expert consultation | Item did not adequately match the factor label |
| INS02 | 67) I can engage in dialogue with clients/colleagues about technology | Expert consultation | Item did not adequately match the factor label |
| INS03 | 68) I can facilitate the dialogue about technology among clients/colleagues | Expert consultation | Item did not adequately match the factor label |
| INS04 | 69) I can tailor the information I give to what clients/colleagues need to apply technology | Expert consultation | |
| INS05 | 70) I can explain the impact of technology to clients/colleagues on their work | Expert consultation | |
| | **Informing (INF)** | | |
| INF01 | 71) I inform clients/colleagues about the usefulness of technology | Expert consultation | Item did not adequately match the factor label |

| | | | |
|---|---|---|---|
| INF02 | 72) I advise clients/colleagues on the use of technology | Expert consultation | Multicollinearity ($r_s$ > 0.7) |
| INF03 | 73) I inspire clients/colleagues to use technology | Expert consultation | |
| INF04 | 74) I encourage clients/colleagues to be alert to the safe use of technology | students' digital competence scale (SDiCoS) [20] | |
| | **Leading (LEA)** | | |
| LEA01 | 75) Colleagues and clients come to me for advice and questions about (new) technology | TRAM [15,16] | |
| LEA02 | 76) My supervisor asks me for advice on (new) technological developments | TRAM [15,16] | |
| LEA03 | 77) I find it difficult to figure out how (new) technology works on my own | TRAM [15,16] | Item was negatively formulated |

| | | | |
|---|---|---|---|
| LEA04 | 78) I have expertise in implementing (new) technology | Expert consultation | |
| LEA05 | 79) I only use (new) technology if I see that others are using it too | Extended UTAUT [14] | Item was negatively formulated |
| **Deepening (DEE)** | | | |
| DEE01 | 80) I actively seek substantiation to deploy a (new) technology | Expert consultation | |
| DEE02 | 81) I don’t really know what the possibilities of technology are | Questionnaire to assess digital competencies and needs of educating nurses [43] | Item was negatively formulated |
| DEE03 | 82) I can assess whether technological innovations are appropriate for my professional practice | Expert consultation | |
| **Connecting (CON)** | | | |

| | | |
|---|---|---|
| CON01 | 83) I identify the needs and requirements of colleagues and clients regarding technology knowledge and skills | Expert consultation |
| CON02 | 84) I interact with other professionals in the field who use technology in their work | Expert consultation |
| CON03 | 85) Through my connections with the other professionals/suppliers in the practice, I remain well informed about technology used there | Expert consultation |
| CON04 | 86) I advise my organisation solicited and unsolicited on the use of (new) technology by clients/colleagues | Expert consultation |
| **Improving (IP)** | | |

| | | | |
|---|---|---|---|
| IP01 | 87) I have ideas on how to improve the use of technology in treatment and counselling | Expert consultation | |
| IP02 | 88) I have ideas about how technology can be improved to better suit the profession of addiction care professionals | Expert consultation | Multicollinearity ($r_s > 0.7$) |
| | **Replacing (REP)** | | |
| REP01 | 89) I advise my organisation about technology that is outdated and needs to be replaced | Expert consultation | Item did not adequately match the factor label |
| REP02 | 90) I advise my organisation about new insights regarding the technology being used | Expert consultation | Multicollinearity ($r_s > 0.7$) |

[a]Items in gray shaded cells have been removed after several steps of item reduction analyses
