## Supplemental Table 2 for "Development of the Technology Readiness Inventory for Professionals (TRIP): field testing in Dutch addiction care professionals"

### S2 Table. Description of technology competencies[a] of health care professionals

| | | |
|---|---|---|
| Basic competencies | Changing (technology) | Improving the care process using innovative technology based on the latest insights and adjusting actions based on self-reflection. |
| | Finding (technology) | Finding and selecting technology using quality criteria that matches client's needs and context. |
| | Being confident (in using technology) | Having confidence in professional's own ability and in the usefulness, added value, effectiveness and safety of technology. |
| | Skilfully using technology | Using technology skilfully, safely, appropriately, competently and (ethically) sensitively. |
| | Instructing your client (about technology) | Instructing patients and their relatives on the use and usefulness of technology. |
| Advanced competencies | Leading | Leading the team and organisation in initiating and implementing technology. |
| | Deepening | Actively exploring and critically assessing technological advancements, applying them to specific requests and their professional context while adhering to ethical and professional standards. |
| | Connecting | Identifying technological needs of colleagues and patients, advising on development and implementation, and interacting with suppliers. |
| | Informing | Educating colleagues on the usefulness and advising and inspiring them to use technology |

| | | |
|---|---|---|
| | Improving | Leveraging on expertise to improve, develop and implement technology that ensures quality of care and well-being. |
| | Replacing | Weighing interests and deciding on the basis of critical reflection and reliable sources to replace technology if necessary. |
| Overarching skill for all professionals | Ethical Reflection | Responsibly choosing and deploying technology by considering ethical, privacy, security, and legal aspects, consulting with stakeholders to weigh various perspectives |

[a]Competencies from the Dutch technology competency model by Den Ouden et al. [18]
